# Comparison of the structure-function properties of wild-type human apoA-V and a C-terminal truncation associated with elevated plasma triglycerides

**DOI:** 10.1101/2023.02.21.23286268

**Authors:** Sylvia Stankov, Cecilia Vitali, Joseph Park, David Nguyen, Leland Mayne, S. Walter Englander, Regeneron Genetics Center, Michael G. Levin, Marijana Vujkovic, Nicholas J. Hand, Michael C. Phillips, Daniel J. Rader

## Abstract

**Background:** Plasma triglycerides (TGs) are causally associated with coronary artery disease and acute pancreatitis. Apolipoprotein A-V (apoA-V, gene *APOA5*) is a liver-secreted protein that is carried on triglyceride-rich lipoproteins and promotes the enzymatic activity of lipoprotein lipase (LPL), thereby reducing TG levels. Little is known about apoA-V structure-function; naturally occurring human *APOA5* variants can provide novel insights.

**Methods:** We used hydrogen-deuterium exchange mass spectrometry to determine the secondary structure of human apoA-V in lipid-free and lipid-associated conditions and identified a C-terminal hydrophobic face. Then, we used genomic data in the Penn Medicine Biobank to identify a rare variant, Q252X, predicted to specifically eliminate this region. We interrogated the function of apoA-V Q252X using recombinant protein *in vitro* and *in vivo* in *apoa5* knockout mice.

**Results:** Human apoA-V Q252X carriers exhibited elevated plasma TG levels consistent with loss of function. *Apoa5* knockout mice injected with AAV vectors expressing wildtype and variant *APOA5*-AAV recapitulated this phenotype. Part of the loss of function is due to reduced mRNA expression. Functionally, recombinant apoA-V Q252X was more readily soluble in aqueous solutions and more exchangeable with lipoproteins than WT apoA-V. Despite lacking the C-terminal hydrophobic region (a putative lipid binding domain) this protein also decreased plasma TG *in vivo*.

**Conclusions:** Deletion of apoA-V’s C-terminus leads to reduced apoA-V bioavailability *in vivo* and higher TG levels. However, the C-terminus is not required for lipoprotein binding or enhancement of intravascular lipolytic activity. WT apoA-V is highly prone to aggregation, and this property is markedly reduced in recombinant apoA-V lacking the C-terminus.

## Introduction

Plasma triglyceride (TG) levels are causally associated with coronary artery disease (CAD)^1, 2^ and hypertriglyceridemia (hyperTG)-induced acute pancreatitis^3^. Approximately 10% of US adults have hyperTG^4^. However, current approaches to reduce to TGs (statins, fibrates, and omega 3 fatty acid products) are often insufficient in reducing extremely elevated TGs,^4, 5^ preventing pancreatitis, or reducing cardiovascular disease risk, and thus elevated TGs represents an insufficiently met medical need^4, 6^.

Human genetics studies can be leveraged to identify novel TG-lowering therapeutic axes. Loss-of-function (LoF) variants in apolipoprotein C-III (apoC-III, gene *APOC3*) are associated with reduced TG levels and protection from CAD^7–9^. Careful structure-function analysis of these variants has led to the development of several approaches targeting *APOC3*, including an *APOC3* antisense oligonucleotide for treatment of familial chylomicronemia to prevent pancreatitis^10^. This work demonstrates that study of TG and TG-rich lipoprotein (TRL) metabolism via human genetics provides valuable insight towards mechanisms that can reduce hyperTG, risk of pancreatitis, and potentially risk of cardiovascular disease.

Apolipoprotein A-V (apoA-V, gene *APOA5*) is a potent modulator of TG metabolism through its ability to promote the activity of lipoprotein lipase (LPL)^11–14^. ApoA-V is secreted from the liver and associates with TRLs in the blood, which present it to the LPL- glycosylphosphatidylinositol anchored high density lipoprotein binding protein 1 (GPIHBP1) complex on the surface of endothelial cells. ApoA-V is remarkably effective in decreasing plasma TG levels despite its low absolute concentration in human plasma (∼150 ng/ml)^15^. LoF mutations in *APOA5* have been associated with elevated TGs^16–18^, increased risk of hyperTG-induced acute pancreatitis^19–21^, and increased risk of CAD^22–24^. Recently, work by Chen et al. and Balasubramaniam et al. demonstrated that apoA-V associates with a neo-epitope on the angiopoietin-like-protein 3/8 (ANGPTL3/8) complex, competing with LPL for that binding site and blocking ANGPTL3/8-mediated LPL inhibition^25, 26^. ApoA-V has also been shown to participate in uptake of remnant particles and may play a role in hepatocyte very low-density lipoprotein (VLDL) secretion^13, 27–30^. Despite this work, little is known about apoA-V structure and its relationship to function.

We mapped the complete secondary structure of recombinant wild-type (WT) apoA-V in both the lipid-free and lipid-associated states using hydrogen-deuterium exchange mass spectrometry (HX MS) and identified a highly amphipathic helical region in the C-terminus. Mining the Penn Medicine Biobank (PMBB), we identified carriers of apoA-V Q252X (rs149808404), a very rare naturally-occurring genetic variant that bears a termination codon which results in elimination of the C-terminal hydrophobic face. Carriers of this variant have elevated plasma TG levels, a phenotype which is recapitulated *in vivo* in *apoa5* knockout (KO) mice expressing apoA-V Q252X. Surprisingly, functional assessments of recombinant apoA-V Q252X reveal this variant’s ability to exchange onto lipoproteins and to reduce plasma TG when injected intravascularly. Our work demonstrates that apoA-V truncation at residue 252 eliminates the C-terminal hydrophobic region and reduces the production of apoA-V by the liver. However, the truncated protein itself is effective in promoting LPL-mediated lipolysis and is much less prone to self-aggregation compared with WT apoA-V.

## Methods

### Recombinant apoA-V

Recombinant WT and Q252X human apoA-V were obtained from DNA-protein Technologies LLC in Oklahoma City (www.DNA-Protein.com)^31,32^ or the laboratory of W. Sean Davidson at the University of Cincinnati^33^. Proteins were solubilized according to the method described by Castleberry et al.^33^ into a final buffer of 10mM NH4HCO3 with 5mM DTT, pH7.8. Protein concentration was determined using Pierce Microplate BCA Protein Assay Kit – Reducing Agent Compatible (Thermo Fisher, Waltham, MA).

For HX MS, apoA-V protein was freshly dialyzed from 6M GdnHCl with 10mM DTT into 50mM sodium citrate buffer (pH 3.8) at 4°C before use. Protein concentration was determined by absorbance at 280 nm. Small unilamellar vesicles (SUV, ∼20nm diameter) of dimyristoyl phosphatidylcholine (DMPC) (Avanti Polar Lipids, Alabaster AL) at 10mg/ml in aqueous buffer were prepared by sonication^34^.

### Circular dichroism measurements

The average *α*-helix content of apoA-V was determined by circular dichroism spectra at room temperature using a Jasco J-810 spectropolarimeter^32^. The spectra were analyzed as previously described to obtain the *α*-helix contents^35, 36^.

### Hydrogen-deuterium exchange mass spectrometry

HX MS methods were applied as previously described^36–38^. The locations, stabilities, and dynamics of *α*-helical segments within the apoA-V protein were assessed under lipid-free and lipid-associated conditions. Briefly, HX was initiated by diluting prepared apoA-V into D2O buffer to a final concentration of about 0.05 mg/ml. The HX kinetics for amides at known positions throughout the protein were determined by fragment-separation method followed by MS analysis^36, 37, 39^. Time-dependent hydrogen to deuterium exchange was measured for each peptide fragment at pH 3.8 and 25°C. Exchange was quenched by lowering pH to 2.5 and temperature to 0°C, followed by proteolysis, high-performance liquid chromatography (HPLC) separation of peptides, and peptide mass and sequence analysis by LTQ Orbitrap XL mass spectrometer (Thermo Fisher Scientific).

Since secondary structure in apoA-V is limited to *α*-helix or helical bundles, protected amides indicate helix location^40, 41^. The degree of HX protection measures helix stability. The observed HX rate curve was compared to a reference rate curve calculated assuming the fragment is in a disordered, random coil state where amide hydrogens are unprotected from exchange with water^42, 43^. The protection factor (Pf, reference rate/observed rate) was used to calculate the free energy of HX (ΔGHX = RT ln Pf) helix unfolding^36, 37^.

### Identification of carriers of APOA5 variants in PMBB, plasma measurements, and FPLC

Plasma samples were obtained from individuals enrolled in the PMBB. Patients participating in the PMBB are consented for the storage of biological specimens, genetic sequencing, and access to all available electronic health record data. The study was approved by the Institutional Review Board of the University of Pennsylvania. *APOA5* variant carriers and controls were identified from a subset of individuals in the PMBB who had exome sequencing (N=41,759). Non-carriers were matched by age, sex, ancestry, BMI, and history of diabetes mellitus diagnosis and screened to exclude individuals carrying any nonsynonymous coding variants in *APOA*5 and *APOC3*. N=73 apoA-V G162C (rs2075291) variant carriers were matched 1:1 to non-carriers (N=73). N=13 apoA-V Q252X (rs149808404) variant carriers were matched 1:3 to non-carriers (N=39). Fasting status of PMBB donors is unknown at time of blood draw.

Carrier and non-carrier plasma samples were assayed for TG, total cholesterol, low-density lipoprotein cholesterol (LDL-C), non-high-density lipoprotein (non-HDL-C), high-density lipoprotein (HDL-C), free cholesterol, phospholipids, apoA-I, apoB, apoC-II, and apoC-III using an Axcel clinical autoanalyzer (Alfa Wassermann Diagnostic Technologies). The association of plasma lipid and apolipoprotein concentrations or ratios with carrier status was determined using a multivariate linear regression analysis adjusted for age, sex, ancestry, BMI and history of diabetes mellitus diagnosis as covariables. Statistical analysis was performed using R (version 4.2.1)^44^.

N=5 representative carrier and non-carrier plasma samples were pooled for separation by fast protein liquid chromatography (FPLC) on a Superose 6 gel-filtration column (GE Healthcare Life Sciences) into 500μl fractions. TG and cholesterol content in 100μl of each fraction was assayed using Infinity Liquid Stable triglyceride or cholesterol reagent (Thermo Fisher Scientific) in 96-well microplates then read with Synergy Multi-Mode Microplate Reader (BioTek).

### AlphaFold predictions of WT and Q252X apoA-V structure

We used AlphaFold2 computational methods to predict the protein structures of WT and Q252X apoA-V^45, 46^. The reference sequence for full length human apoA-V was obtained from Uniprot^47^ (accession number Q6Q788). For structure prediction we used the sequence corresponding to the mature protein, which corresponds to the full-length sequence after removing the signal peptide (aa 1-23). The structure prediction for *APOA5*-*Q275X* (mature protein, Q252X) was obtained from the mature apoA-V protein sequence by removing aa 252-343. Predictions were made using the Colab AlphaFold2 notebook with MMseqs2 (48 recycles)^45, 48^. Structure visualization and analysis was performed using UCSF ChimeraX 1.4 (Resource for Biocomputing, Visualization, and Informatics, University of California, San Francisco)^49, 50^.

### MVP

The VA Million Veteran Program (MVP) is a longitudinal genomic and precision medicine study which recruits participants receiving care from the Veterans Health Administration. Details of MVP have been previously described^51^. More than 900,000 participants have been recruited, with genome-wide genotyping performed on approximately 650,000 participants. Participants were genotyped on the MVP 1.0 custom Axiom array (Thermo Fisher Scientific). Details of genotype quality control have been previously described^52^. After standard quality control, genotypes were imputed to the TOPMed reference panel^53^. Genetic ancestry of participants was determined using HARE^54^. Related individuals were removed using KING^55^.

Prevalent levels of circulating lipids (TG and HDL-C) were obtained by querying the MVP database for clinically obtained measurements in the Veterans Health Administration electronic health record system. Associations between rs2075291, rs149808404, and lipid measurements were performed among participants of each genetic ancestry, accounting for age, sex, and 10 genetic principal components. Linear regression was performed to test associations with lipid levels.

### UKB

Publicly available UK Biobank (UKB, application 26041 and 48511) exome-based association statistics were accessed on Genebass (https://app.genebass.org/)^56^. TG, apoB, HDL-C and apoA-I estimates were obtained by querying each variant using Variant PheWAS. For G162C, alternate allele count = 135, allele number = 735926 and alternate allele frequency = 0.00018. For Q252X, alternate allele count = 72, allele number = 735924, and alternate allele frequency = 0.00010.

### GLGC

Publicly available Global Lipids Genetics Consortium (GLGC)^57–59^ results files were accessed at https://csg.sph.umich.edu/willer/public/glgc-lipids2021/. “Trans-ancestry GWAS summary statistics for HDL, LDL, etc.” were queried for TG and HDL-C associations. METAL meta-analysis results are shown.

### Cloning and production of WT and variant AAVs

Human *APOA5* cDNA was purchased from the DNASU Plasmid Repository (HsCD00719454). A 1.14kb band was amplified using Q5 High-Fidelity DNA polymerase (New England Biolabs (NEB), Ipswich, MA, M0491S) and primers F: 5’- AATAAACTCGAGCCGCCACCATGGCAAGCATGGCTG-3’ and R: 5’- ATTATAGAATTCTCAGGGGTCCCCCAGATGGCTGTG-3’, digested with XhoI (NEB, R0146S) and EcoRI-HF (NEB, R3101S) then sub-cloned into dephosphorylated XhoI and EcoRI-HF digested pcDNA3.1/myc-His(-)A expression plasmid (Invitrogen, Waltham, Massachusetts, V80020). Note, the cloned open reading frame is not tagged. Insert sequence was confirmed by Sanger sequencing.

To generate the Q252X mutation, Integrated DNA Technologies (IDT, Coralville, IA) gBlocks Gene Fragment 5’-AATAATGGCCGGCCCGGACCCCTAGATGCTCTCCGAGGAGGTGCGCCAGCGACTTCAGGCTTTCCGCCAGGACACCTACCTGCAGATAGCTGCCTTCACTCGCGCCATCGACCAGGAGACTGAGGAGGTCCAGCAGCAGCTGGCGCCACCTCCACCAGGCCACAGTGCCTTCGCCCCAGAGTTTCAACAAACAGACAGTGGCAAGGTTCTGAGCAAGCTGCAGGCCCGTCTGGATGACCTGTGGGAAGACATCACTCACAGCCTTCATGACCAGGGCCACAGCCATCTGGGGGACCCCTGATAATGAATTCCATATAT-3’ was digested with FseI (NEB, R0588S) and EcoRI-HF then purified (QIAquick PCR Purification Kit, 28104). This fragment was sub-cloned into FseI and EcoRI-HF cut WT h*APOA5* pcDNA generated above. The insert sequence was confirmed by Sanger sequencing. Note, the Q252X construct is identical to the WT construct except for the introduction of the single nucleotide polymorphism observed in carriers (Gln CAG à *Stop TAG).

Both pcDNA constructs were digested with NheI-HF (NEB, R3131S) and SpeI-HF (NEB, R3133S). The respective inserts were ligated into pENN.AAV.TBG.PI.mcs.rBG vector digested with SpeI and colonies were screened by EcoRI-HF digestion for correct orientation. The AAV serotype 8 vector plasmid containing the liver-specific thyroxine-binding globulin (TBG) promoter was provided by the Vector Core of the University of Pennsylvania^60^. Insert sequence was confirmed by Sanger sequencing and vector was confirmed by restriction digest. AAV production, purification, and titration from both cloned *APOA5* and the Null (empty) vector plasmids were performed by the Vector Core of the University of Pennsylvania.

### AAV expression in mice

*Apoa5* KO mice were provided by Robert O. Ryan (University of Nevada, Reno) and bred at the University of Pennsylvania^11, 61^. Mice were housed in a small animal facility on a 12-hour light/dark cycle (lights off from 7:00pm to 7:00am), fed with standard chow diet, and provided access to water ad libitum. All animal experiment procedures were reviewed and approved by the Institutional Animal Care and Use Committees of the University of Pennsylvania.

Prior to AAV injection, mice were fasted 4 hours then bled via retro-orbital sinus using heparin-coated glass tubes while under isoflurane-induced anesthesia. Plasma was separated by centrifugation of blood at 10,000 x g for 7 minutes at 4°C. Plasma TG levels were determined using Axcel autoanalyzer (Alfa Wassermann) and used to assign experimental groups with similar starting plasma TG levels (week 0, baseline).

Male mice approximately 8 to 18 weeks of age (age-matched within study) were administered 3E11 or 3E12 vector genomes (vg)/mouse Null AAV or WT or Q252X h*APOA5*-AAV via intraperitoneal injection. Female mice 19 weeks of age were administered 3E12 vg/mouse WT or Q252X h*APOA5*-AAV via intraperitoneal injection. AAVs were diluted in sterile PBS under a sterile chemical hood and administered using insulin syringes. Blood was collected at indicated time points following a 4 hour fast via retro-orbital bleeding, and plasma was separated as described.

### Lipid measurements and FPLC of mouse plasma

Plasma TG and total cholesterol measurements were performed on individual mouse plasma samples using an Axcel autoanalyzer (Alfa Wassermann Diagnostic Technologies).

Plasma samples were pooled by experimental group for separation by FPLC and assayed for TG and cholesterol content as described.

### Liver TG measurements

At time of sacrifice, mice were perfused with ice cold PBS and their livers snap frozen in liquid nitrogen then stored at -80°C. To assay TG content, 150-250mg liver tissue was homogenized in 3x volumes PBS and shaken on a Tissue Lyzer for 2 minutes at 30Hz. Homogenate was immediately diluted 1:5 in PBS and 20μl added in duplicate to 96-well microplates. 20μl of 1% deoxycholate solution was added and allowed to incubate at 37°C for 5 minutes. 200μl of Infinity Liquid Stable triglyceride reagent (Thermo Fisher Scientific) was added to each well and allowed to incubate at 37°C for 30 minutes before reading with Synergy Multi- Mode Microplate Reader (BioTek). For normalization, the same 1:5 diluted homogenates were assayed for protein content using Pierce BCA protein assay kit (23227, Thermo Scientific, Waltham, MA).

### Liver gene expression

At time of sacrifice, mice were perfused with ice cold PBS and their livers snap frozen in liquid nitrogen then stored at -80°C. For whole liver RNA, tissue was homogenized directly in RNAzol reagent (R4533-100ML, Sigma-Aldrich, St. Louis, MO) according to the manufacturer’s protocol. For nuclear RNA, approximately 50mg of liver tissue was gently homogenized with Dounce Homogenizer in 1mL chilled lysis buffer (10mM Tris-HCl, 10mM NaCl, 3mM CaCl2, 0.1% NP-40, 0.1% Tween-20, 5μl/mL Dnase, 2μl/mL Rnase-I, 1x PBS + 1% BSA). After incubation on ice, samples were pipetted through a 100μm filter on top of a 30μm filter, followed by 1ml wash buffer (1x PBS (Mg-, Cl-), 1% BSA). Filtered samples were centrifuged at 500 x g for 5 minutes at 4°C to pellet nuclei. After discarding supernatant, pellets were resuspended in 1ml wash buffer followed by centrifugation, repeating until there were no visible clumps or debris. After final centrifugation, nuclei were re-suspended in 500μl RNAzol reagent and RNA extracted according to the manufacturer’s protocol.

1μg of RNA was treated with RQ1 Rnase-free Dnase (Promega, Madison, WI) then reverse transcribed using the High-Capacity cDNA Reverse Transcription kit (Thermo Fisher Scientific). Gene expression levels of human *APOA5*, murine *Rpl30*, murine *B2m* and murine *Hmbs* were determined by quantitative real-time PCR using a QuantStudio 7 Real-Time PCR System. Human *APOA5* gene expression was normalized to the geometric mean Ct of the murine *Rpl30*, *B2m* and *Hmbs* gene expression from the same sample. Relative fold expression is in comparison to mean of WT *APOA5*. Primer sequences used for these reactions:

- *APOA5*: 5’ –TGGCTCTTCTTTCAGCGTTT– 3’, 5’ –TTCTGCTGATGGATCTGCTC– 3’
- *Rpl30*: 5’ –ATGGTGGCCGCAAAGAAGACGAA– 3’, 5’ - CCTCAAAGCTGGACAGTTGTTGGCA– 3’
- *B2m*: 5’ -CATGGCTCGCTCGGTGAC- 3’, 5’ -CAGTTCAGTATGTTCGGCTTCC- 3’
- *Hmbs*: 5’ –ATGAGGGTGATTCGAGTGGG- 3’, 5’ -TTGTCTCCCGTGGTGGACATA- 3’

### mRNA Stability time course

CHO-K1 cells (ATCC, CCL-61, Masassas, VA) were seeded in T75 flasks and maintained in Ham’s F-12K (Kaighn’s) Medium (Gibco 21127022, Waltham, MA) supplemented with 10% Fetal Bovine Serum (Cytiva, SH30910.03, Marlborough, MA). Each flask was transfected with 20μg WT h*APOA5* pcDNA or Q252X h*APOA5* pcDNA using Lipofectamine 3000 Transfection Reagent (Invitrogen, L3000015, Waltham MA) per manufacturer’s protocol. 24 hours later, cells were re-plated into 12-well plates at 2x10^5^ cells per well. 24 hours later, media was aspirated from hour 0 (baseline) wells and cells were collected in 500μl RNAzol reagent (R4533-100ML, Sigma-Aldrich, St. Louis, MO) and stored at -80°C. Then, 1mL fresh media was replaced in each remaining well, followed by dropwise addition of 120μl of 0.5mg/ml actinomycin D (Sigma-Aldrich, A9415-5MG, St. Louis, MO). Media was aspirated and cells collected as described at 2-hour intervals over 12 hours.

RNA was extracted according to manufacturer’s protocol. 0.5μg of RNA was reverse transcribed using the High-Capacity cDNA Reverse Transcription kit (Thermo Fisher Scientific). Gene expression levels of human *APOA5*, hamster *GNB1*, hamster *RPS16*, and hamster *EIF3K* were determined by quantitative real-time PCR using a QuantStudio 7 Real-Time PCR System. Human *APOA5* gene expression was normalized to the geometric mean Ct of the hamster *GNB1*, *RPS16*, and *EIF3K* gene expression from the same sample. Relative fold expression in comparison to mean of WT *APOA5* at time 0. Primer sequences used for these reactions:

- *APOA5*: 5’ –TGGCTCTTCTTTCAGCGTTT– 3’, 5’ –TTCTGCTGATGGATCTGCTC– 3’
- *Gnb1*: 5’ -CCATATGTTTCTTTCCCAATGGC- 3’, 5’ -AAGTCGTCGTACCCAGCAAG- 3’
- *Rps16*: 5’ -TGAAGGGTGGTGGTCATGTG- 3’, 5’ -TCAGCTACAAGCAGGGTTCG- 3’
- *EIF3K*: 5’ -AGCCCAAAAACATCGTGGAGA- 3’, 5’ – TCATTGAGGAAGGGGCAGAAG- 3’

### Western blotting

Samples were combined with buffer (0.125M Tris HCl, pH6.8, 4% SDS, 20% glycerol, 1% 2-Mercaptoethanol, 5% water, bromophenol blue for color) then heated to 95°C for 10 minutes before storage at -80°C. Prepared samples were separated by SDS-PAGE using the NuPage system (Invitrogen, Waltham, MA). Recombinant apoA-V Q252X protein could not be detected with CST mAb #3335, Abcam ab55977, Invitrogen MA1-16809, OriGene TA507164, or Sigma-Aldrich ABS443. Instead, plasma from mice injected with 3E12 vg Q252X h*APOA5*-AAV was used as primary antibody. When diluted 1:100, this plasma can detect recombinant apoA-V Q252X protein **(Figure S8)**. IRDye 680 donkey anti-mouse IgG secondary antibody (926-68072, LI-COR, Lincoln, NE) was used at dilution 1:10,000. Membranes were scanned using the Odyssey Fc Imaging System (LI-COR, Lincoln, NE).

### 125I radiolabeling

WT or Q252X apoA-V protein (10mM NH4HCO3, 5mM DTT, pH7.8) was iodinated with ^125^I directly using the iodine mono-chloride method and as previously described^60, 62^. Approximately 0.2mg free protein in 2.0ml was iodinated with 0.5mCi of ^125^I (PerkinElmer). Proteins were combined with 300μl of 1M glycine, and 150μl of 1.84M NaCl/2.64μM ICl solution, vortexed, and applied to a PG-10 desalting column (Amersham Biosciences) that was pre-equilibrated with 0.15M NaCl/1mM EDTA solution. Iodinated proteins were eluted to a final volume of 2.5ml in NaCl/EDTA solution, transferred to 2000 MWCO Slide-A-Lyzer dialysis cassettes (66203, Thermo Scientific, Waltham, MA) and dialyzed against PBS. Protein concentration was measured with Pierce BCA protein assay and ^125^I activity measured by gamma counting.

### 125I protein incubation with plasma followed by FPLC

0.5μg ^125^I WT or Q252X apoA-V protein was brought to 100μl with PBS and incubated with 225μl *apoa5* KO plasma or human plasma. Mixtures were incubated in a water bath at 37°C for 3 hours before separation by FPLC. ^125^I activity was measured in 100μl of each fraction and expressed as percent of total eluted counts. TG and cholesterol content in 100μl of each fraction was assayed using Infinity Liquid Stable triglyceride or cholesterol reagent (Thermo Fisher Scientific) in 96-well microplates then read with Synergy Multi-Mode Microplate Reader (BioTek).

### rHDL formulation

Synthetic HDL containing recombinant human WT or Q252X apoA-V (rHDL) were prepared with egg L-*α*-phosphatidylcholine (PC) (egg, chicken PC, 840051C-200mg, Avanti Polar Lipids, Alabaster, AL). Egg PC solution in chloroform was dried under nitrogen, without heat in glass tubes (14-961-29, Fisher Scientific, Waltham, MA). PBS was added and tubes vortexed on high for 1 minute, followed by addition of sodium cholate in PBS (cholic acid sodium salt, 229101, Millipore, Burlington, MA, ∼11.4mM in final volume). Tubes were vortexed again on high for 1 minute before incubation in a water bath at 37°C for 1.5 hours, vortexing every 15 minutes. Respective recombinant apoA-V in 10mM NH4HCO3, 5mM DTT, pH7.8 was added to each tube to achieve a molar ratio of 1 apoA-V to 300 egg PC. Tubes were vortexed for 30 seconds before incubation in a water bath at 37°C for 1 hour. Solutions were transferred to 2000 MWCO Slide-A-Lyzer dialysis cassettes (66203, Thermo Scientific, Waltham, MA) and dialyzed 3 days at 4°C against PBS. Buffer was changed twice daily.

To verify successful incorporation of apoA-V protein into rHDL particles, formulations were fractionated by FPLC. Protein content in each fraction was monitored by measuring the UV absorbance at wavelength 280nm. The UV spectra were evaluated for the presence of a distinct peak in fractions corresponding to lipoprotein-sized particles or lipid-free protein.

### In vivo rHDL studies

9 weeks after 3E12vg/mouse WT or Q252X h*APOA5*-AAV administration, mice were fasted 4 hours and bled for baseline TG measurements, as described. 5μg WT apoA-V rHDL or equivalent moles Q252X apoA-V rHDL (3.35μg) were brought to 150μl with PBS and injected retro-orbitally using insulin syringes. Non-AAV injected *apoa5* KO mice were also fasted 4 hours for baseline TG measurements. 20μg WT apoA-V rHDL or equivalent moles Q252X apoA-V rHDL (13.40μg) were brought to 150μl with PBS and injected retro-orbitally using insulin syringes. Mice were bled over 12 hours and plasma TG content assayed using Infinity Liquid Stable triglyceride reagent (Thermo Fisher Scientific), as described. Results are plotted as raw plasma TG values over time or as percent TG of baseline. Area under the curve (AUC) for each mouse was calculated in Prism GraphPad (GraphPad Software, San Diego, CA www.graphpad.com) using Y=0 as baseline and using data expressed as percent TG of baseline.

### DMPC clearance

ApoA-I was isolated from human plasma using the method described by Vega et al.^63^, with minor adaptations. For DMPC clearance, methods were adapted from Castleberry et al^33^. The DMPC to protein mass ratio (2.5:1) was maintained but the assay was miniaturized to a total volume of 100μl and performed in a 96-well plate. DMPC (Avanti Polar Lipids, Alabaster AL) was dried under nitrogen without heat, in a glass tube, brought to a concentration of 5mg/ml in 10mM NH4HCO3, 5mM DTT, pH7.8, vortexed and briefly sonicated. DMPC was diluted to 1mg/ml immediately before distribution to wells. Buffer, equivalent moles WT apoA-V (17μg) or Q252X apoA-V (11.39μg) or equivalent mass (17μg, matching WT apoA-V) or moles (12.27μg) apoA-I in 10mM NH4HCO3, 5mM DTT, pH7.8 were added to wells. Plates were read at 325nm with Synergy Multi-Mode Microplate Reader (BioTek) every minute for 1 hour. Data expressed as fraction of turbidity remaining (ratio of first reading).

### Statistical analyses

PMBB, MVP, UKBB and GLGC analyses are detailed under their respective methods. All other – data reported as mean with SEM. Statistical comparisons between 2 groups were performed in Prism GraphPad, using two-tailed unpaired t-test, where statistical significance was defined as p less than 0.05 (GraphPad Software, San Diego, CA www.graphpad.com).

## Results

### HX MS reveals a helical apoA-V secondary structure and helical content increases with lipid association

We performed HX MS on mature human WT apoA-V recombinant protein to solve its secondary structure. In the lipid-free state, the amide NH in the N-terminus up to residue 26 exhibit a Pf=1 and exchange at the theoretical rate expected for a dynamically disordered protein **(Figure S1, Table S1**). Thus, this fragment was determined to be unstructured. In contrast, the peptide spanning residues 27-47 contains 7 amide NH with Pf=20. The following peptides at residues 48- 62 and 48-66 are unprotected, indicating that the 7 protected amide NH form an *α*-helix. The amide NH of residue 41 is hydrogen bonded to the main chain carbonyl of residue 37 (i to i-4). Residues 38, 39, and 40 form the first turn of the helix and their amide hydrogens are not protected by hydrogen bond acceptors. Therefore, the first *α*-helix in lipid-free apoA-V extends between residues 37-47.

When apoA-V is associated with DMPC SUV (lipid), the peptide spanning residues 27-47 contains 9 protected amide NH with Pf=70 (**Figure S2, Table S2)**. These values indicate that in the presence of lipid, the first helix of apoA-V is elongated and stabilized. This analysis was performed for the entirety of the mature apoA-V in the lipid-free and lipid-associated states (**Figure 1).** HX MS was also performed with apoA-V associated with DMPC SUV at pH 7 and 5°C (slow exchange) and yielded helix locations similar to those shown in **Figure 1**, along with vesicles prepared with the more fluid palmitoyl-oleoyl PC (POPC), which also gave similar results (data not shown). Measurements with lipid-free apoA-V at pH 7 were not possible because of limited protein solubility and very rapid exchange rates at neutral pH.

**Figure 1.**
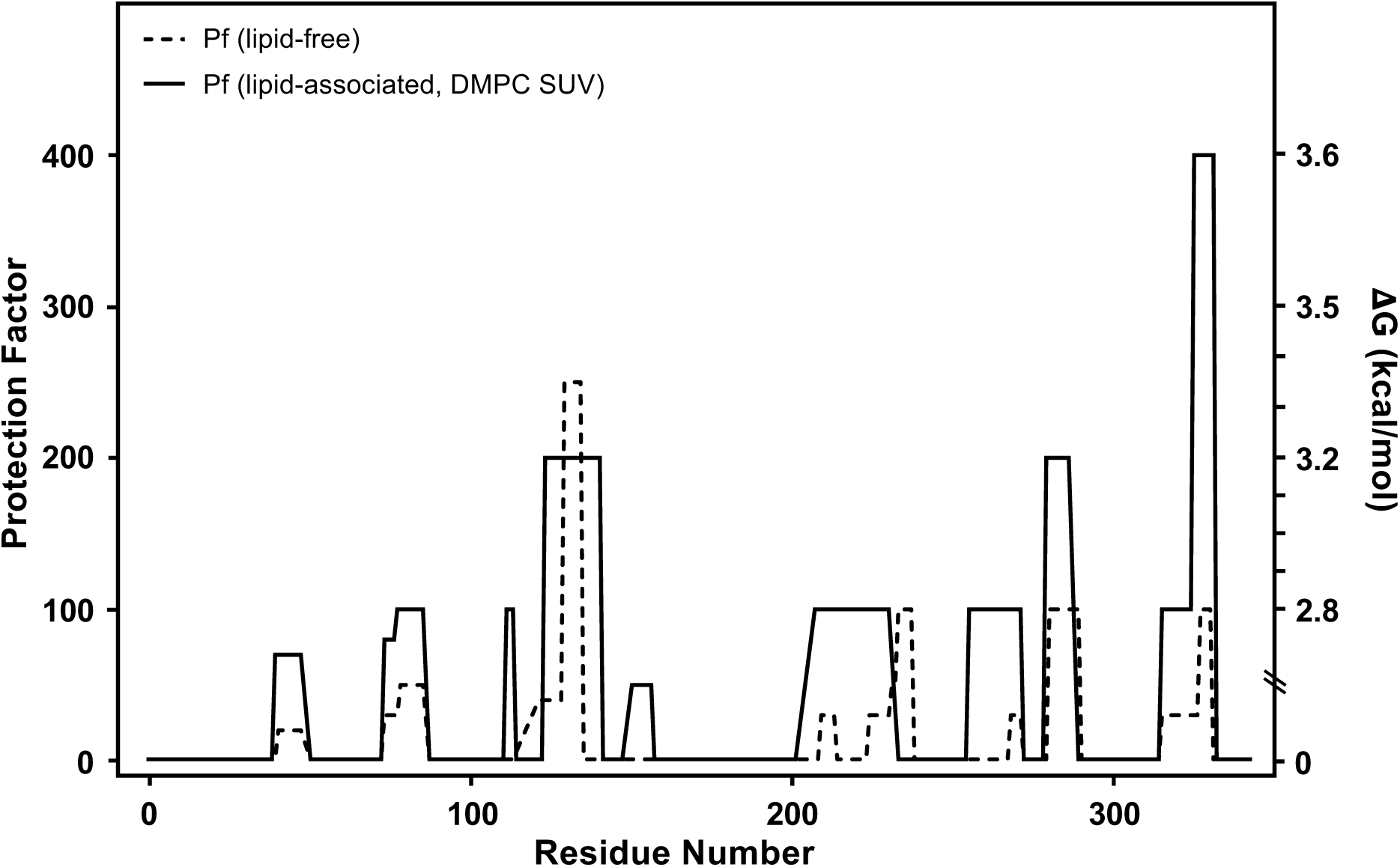
Summary of HX derived α-helix stabilities and secondary structure assignments for human apoA-V in lipid-free and lipid-associated states at pH 3.8 and 25°C. The HX MS kinetic data in Figures S1 and S2 and Tables S1 and S2 were analyzed to obtain Pf values (left Y-axis) for each apoA-V peptide fragment. Pf values for lipid-free apoA-V (dashed line) and lipid- associated apoA-V (solid line) are plotted for the length of apoA-V. The free energy of helix stabilization (*Δ*G) is shown on the right Y-axis. apoA-V, apolipoprotein A5; DMPC, dimyristoyl phosphatidylcholine; HX MS, hydrogen-deuterium exchange mass spectrometry; Pf, protection factor; SUV, small unilamellar vesicles.

Mature apoA-V contains multiple helical segments, most of which are stabilized upon association with DMPC SUV **(Figure 2A, 2B)**. Using circular dichroism measurements, we determined that lipid-free apoA-V has a helix content of ∼35%. Upon association with lipid (DMPC SUV), apoA-V helical content increases to ∼45%, in agreement with the HX results. The increase in helix content is consistent with the fact that amphipathic *α*-helix formation provides a more favorable free energy change that drives binding of exchangeable apolipoproteins to lipid-water interfaces^64^. The Pf values depicted in **Figure 1** indicate that the stabilities of the α-helices in the lipid-free apoA-V molecule are low. Thus, the N-terminal helices have Pf ∼20 which corresponds to a helix stabilization free energy ΔG=1.6 kcal/mol. It follows that these helices are unfolded approximately 5% of the time; the helix opening and closing reaction occurs in a timescale of seconds^37^. The C-terminal helix spanning aa 311-331 which is responsible for initial interaction with the surface of lipoprotein particles^65^ has Pf in the range 20-100 (**Figure 1, S3A**). The ΔG of helix stabilization is 1.6-2.5 kcal/mol indicating that the helix is open about 1-5% of the time. Interaction with lipid increases the Pf to ∼400 and the ΔG of helix stabilization to ∼3 kcal/mol, indicating that the helix is open ∼0.5% of the time.

**Figure 2.**
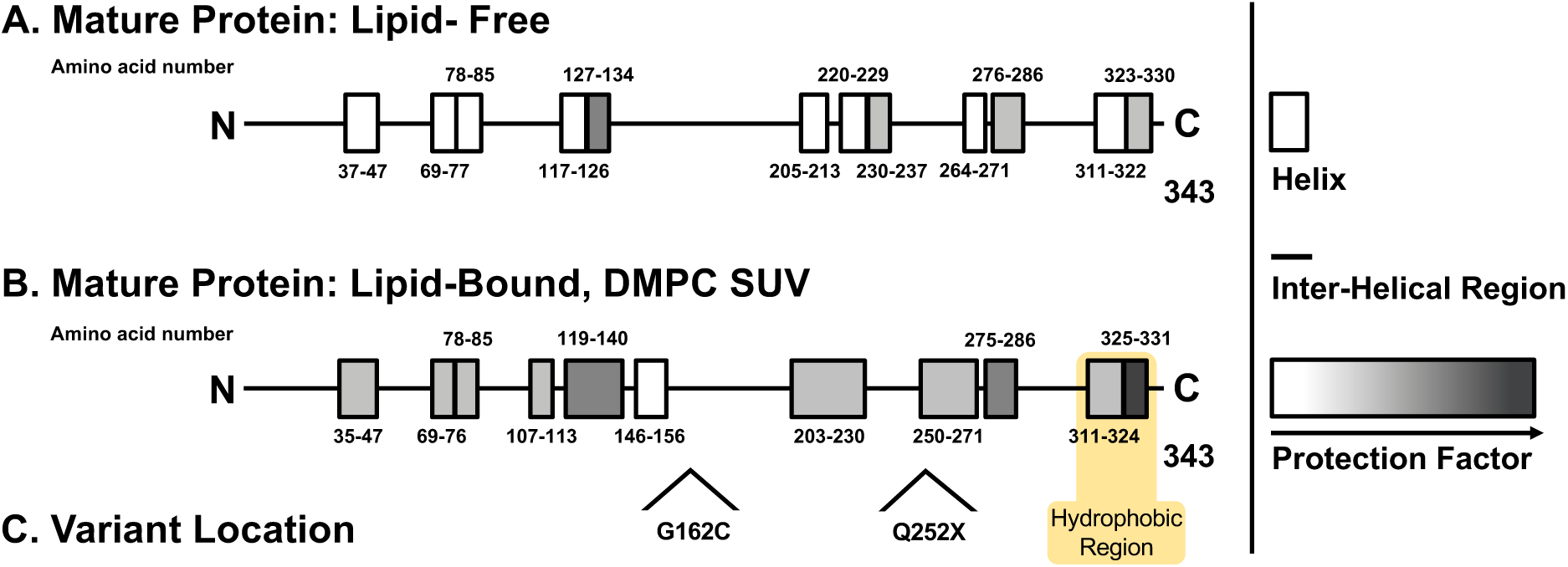
ApoA-V secondary structure determined by HX MS. **A.** HX MS structure for mature apoA-V protein in the lipid-free state (± 2 residues). Rectangles represent ⍺-helices and lines represent disordered structure. Protection against amide HX with water reflects helix stability and is indicated by the shading. **B.** HX MS structure for mature apoA-V protein in the lipid-associated state (± 2 residues). **C.** Locations of variants. C-terminal hydrophobic region is highlighted in yellow. apoA-V, apolipoprotein A5; DMPC, dimyristoyl phosphatidylcholine; HX MS, hydrogen- deuterium exchange mass spectrometry; SUV, small unilamellar vesicles.

We further explored the structural architecture of mature WT apoA-V using Colab AlphaFold2 notebook with MMseqs2^45, 46^. AlphaFold predicts a structure with multiple helical segments, agreeing with our HX MS results **(Figure S4A-C)**. A lack of total agreement between these two methods is reasonable, as HX experimentation is limited to detecting relatively stable helices while AlphaFold predictions include all possible helices, more reflecting the lipid-associated state. Because the N-terminal predictions returned low confidence scores (<70), they were excluded from further analysis. Four helical segments are predicted to pack in a coiled-coil arrangement, to create a previously described helix bundle^41^ (aa 86-139; 143-190; 198-242 and 251-291). Examination of the hydrophobicities and amphiphilicities of these helices suggest that they may form a mild hydrophobic face **(Figure S3A)**. Consistent with our HX MS data, the C-terminal region of apoA-V (aa 310-335) is predicted to form a helical structure with high hydrophobic moment **(Figure S3A-C)**. The clear separation of polar and nonpolar amino acids on the two faces of the helix (**Figure S3B**) as well as the surface map of hydrophobic potential (**Figure S3C**) support the presence of this region. Together, our HX experimental data and AlphaFold predictions suggest that the apoA-V C-terminus forms the primary hydrophobic face. In support, a premature truncation mutant of apoA-V (at residue 292) has markedly decreased lipid and lipoprotein binding ability^40^.

### Humans with the rare truncating apoA-V Q252X variant have elevated plasma TG

We screened the Penn Medicine Biobank (PMBB) for rare naturally-occurring *APOA5* genetic variants associated with elevated TG levels. As a positive control, we identified 73 carriers of apoA-V G162C (rs2075291), a known rare LoF variant (total allele frequency 0.00342 in gnomAD v3.1.2 dataset (GRCh38)^66^) associated with elevated plasma TG^16, 17^, CAD^22, 24^, and hypertriglyceridemia-induced acute pancreatitis^19–21^ **(Table S3)**. Carriers of this variant in the PMBB had significantly elevated plasma TG levels **(Figure 3A)**. In addition, carriers had no significant differences in apoB but elevated TG to apoB ratios when compared to matched non-carriers, a reflection of apoA-V’s specific effect on lipolysis of TG-rich lipoproteins **(Figure 3B, 3C)**.

**Figure 3.**
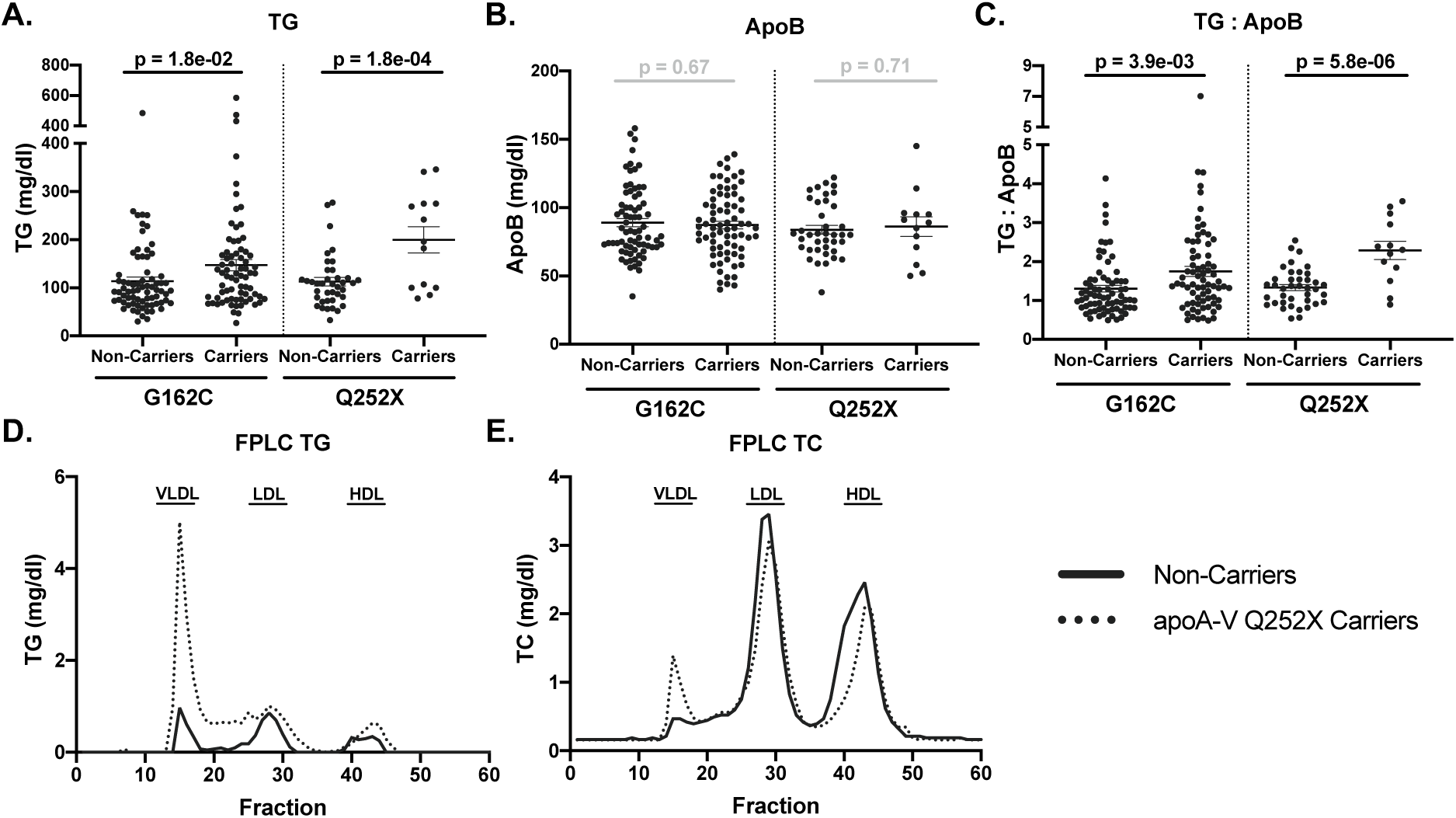
Human apoA-V G162C and Q252X carriers exhibit elevated plasma TG. Variant carriers and matched controls (non-carriers) were identified from exome-wide genotyping in the Penn Medicine Biobank. **A.** Plasma TG. **B.** Plasma apoB. **C.** Plasma TG to apoB ratio. **D.** TG content and **E.** TC content of human plasma lipoproteins, separated by fast protein liquid chromatography (FPLC). Representative PMBB plasma samples were pooled by Q252X variant carrier or matched, non-carrier status prior to fractionation. Fractions corresponding to the elution range of VLDL, LDL and HDL particles are indicated. **A-C.** G162C carriers (heterozygous) N=73, non- carriers N=73. Q252X carriers (heterozygous) N=13, non-carriers N=39. Mean with SEM (brackets). P-value determined by linear regression analysis adjusted by SNP, age, sex, race, BMI and diabetes mellitus status. **D, E.** Line indicates single values. apoA-V, apolipoprotein A5; apoB, apolipoprotein B; BMI, body mass index; FPLC, fast protein liquid chromatography; HDL, high-density lipoprotein; LDL, low-density lipoprotein; SNP, single nucleotide polymorphism; TC, total cholesterol; TG, triglyceride; VLDL, very low-density lipoprotein.

We also identified 13 carriers of a very rare variant apoA-V Q252X (rs149808404; total allele frequency 0.00003 in gnomAD v3.1.2 dataset^66^), a premature truncation variant which is predicted to remove the C-terminal hydrophobic region that we identified **(Figure 2B, 2C, Table S3)**. This variant has an allele frequency of 0.00016 in the PMBB. Compared with matched controls, apoA-V Q252X carriers had significantly elevated plasma TG levels as well as increased TG to apoB ratios with no significant differences in apoB when compared to matched non-carriers **(Figure 3A-C)**. FPLC separation of pooled plasma from representative human apoA-V Q252X carriers and matched, non-carrier controls shows that carriers exhibit a marked increase in VLDL TG and VLDL cholesterol, a modest increase in HDL TG and a modest decrease in LDL and HDL cholesterol **(Figure 3D, 3E).**

Carriers of apoA-V G162C and Q252X show a decrease in HDL cholesterol to apoA-I ratios with no significant differences in apoA-I or HDL cholesterol independently **(Table S4)**. Carriers of both variants show no differences in total cholesterol, LDL cholesterol, non-HDL cholesterol, free cholesterol, and phospholipids **(Table S4)**. ApoA-V Q252X but not G162C carriers show an increase in apoC-II and apoC-III **(Table S4)**.

We mined other biobanks for carriers of this novel apoA-V truncating variant Q252X as well as G162C as a positive control to corroborate the high TG phenotype. In the Million Veteran Program (MVP), carriers of the known LoF variant G162C had significantly elevated plasma TG and decreased HDL-C as expected (**Table S5**). Carriers of apoA-V Q252X also had substantially elevated plasma TG and decreased HDL-C, supporting a LoF phenotype (**Table S5**). In the UK Biobank (UKBB), both G162C and Q252X carriers had elevated plasma TG and decreased HDL-C and apoA-I (**Table S6**). In the Global Lipids Genetics Consortium (GLGC) meta-analysis, apoA-V G162C is associated with elevated plasma TG and decreased HDL-C, but the apoA-V Q252X was not present (**Table S7**).

### ApoA-V Q252X is predicted to be less stable

To investigate the potential effects of the premature stop of translation at position 252, we examined the location of the non-translated amino acid sequence on the AlphaFold predicted WT apoA-V structure **(Figure S4D)** and modeled apoA-V Q252X as a *de novo* protein **(Figure S4E- G)**. The prediction returned an overall lower confidence score compared to what was observed for the WT structure. It is apparent that the truncated variant not only lacks the C-terminal hydrophobic face but also one of the helical regions involved in the formation of the helix bundle. The resulting structure appears to be less stable, consistent with LoF **(Figure S4F)**.

### ApoA-V Q252X has reduced expression, producing a LoF TG phenotype

We used AAV8 vectors to express WT and Q252X apoA-V in the livers of *apoa5* KO mice. Both constructs were identical except for the point mutation Gln (CAG) to *Stop (TAG) at amino acid position 252. Delivery of WT h*APOA5*-AAV showed a dose-dependent decrease in plasma TG, a concomitant decrease in total cholesterol only at the highest dose (3E12), and no effect on body weight over 4 weeks **(Figure S5A-C)**.

For initial phenotype profiling, experiments were performed with 3E11 vg AAV for its specific effect on TGs and not cholesterol. Mice expressing WT apoA-V had a significant ∼60% decrease in plasma TG when compared to mice injected with Null vector **(Figure 4D)**. In contrast, mice expressing apoA-V Q252X exhibit a non-significant (vs. Null) change in plasma TG, which was significantly different when compared to mice expressing WT apoA-V **(Figure 4D)**. FPLC separation of pooled plasma shows a large decrease in VLDL TG in mice expressing WT apoA-V, when compared to mice expressing apoA-V Q252X and Null-injected mice **(Figure 4E).** Mice expressing WT apoA-V exhibit a notable decrease in VLDL cholesterol and concomitant increase in HDL cholesterol **(Figure 4F)**, whereas mice expressing apoA-V Q252X had only a small decrease in VLDL cholesterol and similar HDL cholesterol when compared to Null **(Figure 4F).** AAV injected mice show no significant difference in liver TG content between all groups after four weeks of apoA-V expression **(Figure S6)**. Mice were also administered the higher 3E12 vg AAV dose. Both male and female *apoa5* KO mice expressing apoA-V Q252X exhibit higher plasma TG when compared to littermates expressing WT apoA-V **(Figure S7A, S7B)**.

**Figure 4.**
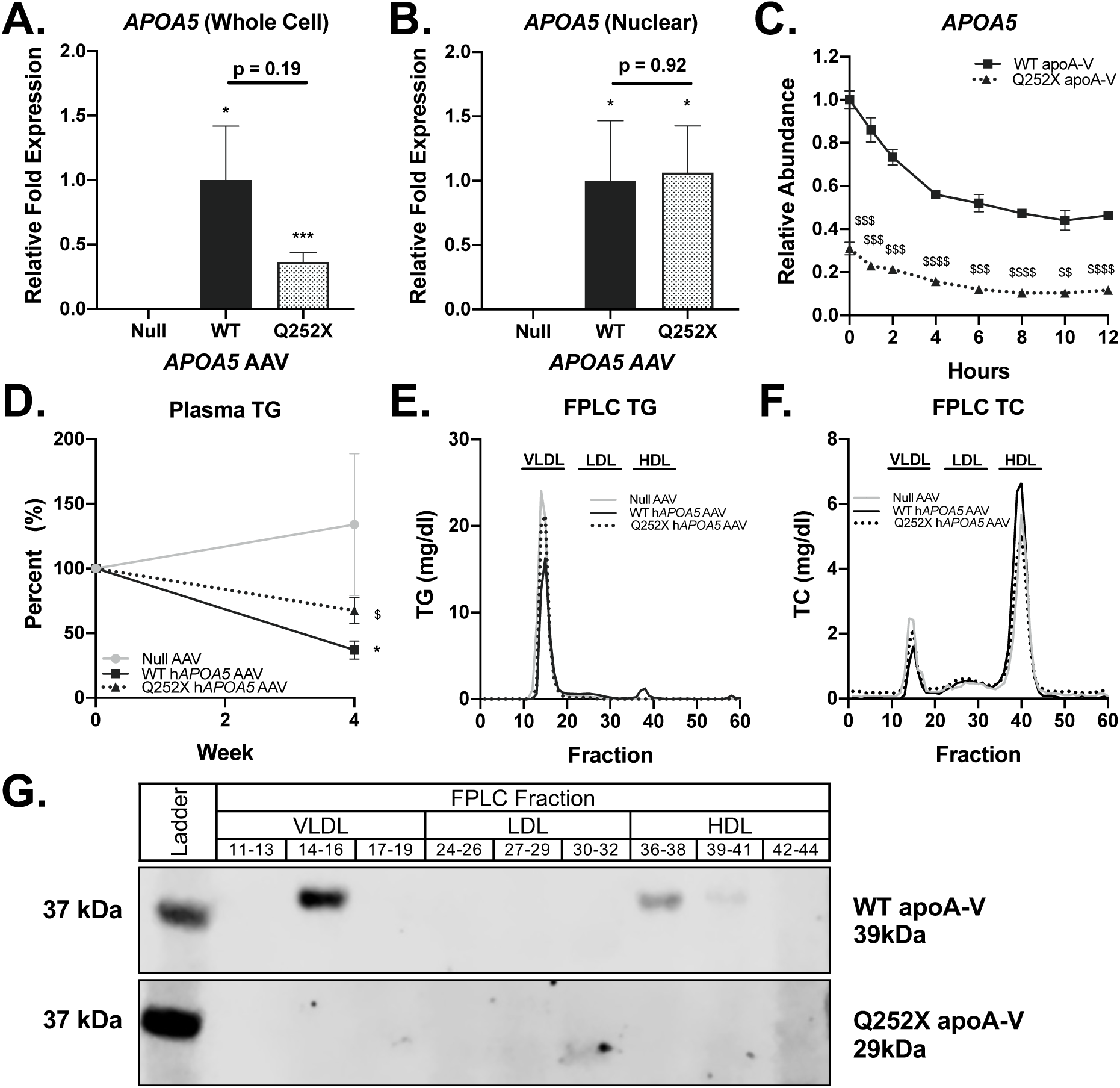
Mice expressing apoA-V Q252X have elevated plasma TG. *Apoa5* KO mice were injected with 3E11 vg Null, WT or Q252X h*APOA5* AAV. **A**. Relative fold expression *APOA5* in whole cell liver lysate (N=4-5). **B**. Relative fold expression *APOA5* in liver nuclear mRNA (N=3- 5). **C**. Relative abundance of *APOA5* over time in CHO cells, transiently transfected to express WT or Q252X *APOA5* then exposed to actinomycin-D. (N=3). **D**. Plasma TG levels at baseline and 4 weeks after AAV administration. Results are expressed as percent of baseline levels (N=5- 11). **E**. TG content and **F**. TC content of plasma lipoproteins, separated by fast protein liquid chromatography (FPLC). Plasma samples were pooled by experimental group prior to fractionation. Fractions corresponding to the elution range of VLDL, LDL and HDL particles are indicated. **G**. ApoA-V content of plasma lipoproteins, separated by fast protein liquid chromatography (FPLC). Indicated FPLC fractions were pooled and immunoblotted to detect WT and Q252X apoA-V. **A-D**. Mean with SEM (brackets). **E, F**. Points indicate single value. Unpaired 2-tailed t-test. p<0.05 = *, p<0.001 = *** vs. Null. p<0.05 = $, p<0.01 = $$, p<0.001 = $$$, p<0.0001 = $$$$ vs. WT. apoA-V, apolipoprotein A5; CHO, Chinese Hamster Ovary; FPLC, fast protein liquid chromatography; HDL, high-density lipoprotein; KO, knockout; LDL, low-density lipoprotein; TC, total cholesterol; TG, triglyceride; VLDL, very low-density lipoprotein.

We then sought to compare expression between WT and Q252X after AAV8 injection. Interestingly, mice expressing apoA-V Q252X had a decrease in hepatic *APOA5* mRNA when compared to mice expressing WT apoA-V, despite receiving the same dose of AAV delivering a cDNA construct **(Figure 4A)**. Isolation of nuclei from these livers reveals similar nuclear WT and Q252X *APOA5* mRNA content (p=0.92), confirming that the groups received identical AAV doses **(Figure 4B)**. This differential expression was reproduced in CHO-K1 cell lines transfected with WT or Q252X apoA-V pcDNA constructs; addition of actinomycin-D to track mRNA stability decreases WT message ∼50% over 12 hours while Q252X message decreases from ∼30% of WT at time 0 to ∼10% of WT by hour 6, where it plateaus through the end of the time course **(Figure 4C)**. Premature stop in the Q252X mRNA may affect stability of the transcript at translation, decreasing mRNA abundance^67, 68^.

We also sought to assess apoA-V protein in plasma lipoprotein fractions. Because no reliable commercially-available ELISA for human apoA-V currently exists, we used antiserum as an antibody to human apoA-V that we confirmed detects both WT and Q252X apoA-V (**Figure S8**). Separation of FPLC fractions by SDS-PAGE and detection by Western blot shows a clear band for WT apoA-V in VLDL fractions with a lighter band in large HDL fractions **(Figure 4G)**. In contrast, apoA-V Q252X appears as only a very faint band in small LDL fractions with no bands in VLDL or HDL, suggestive of substantially decreased truncated protein in plasma **(Figure 4G)**. Overall, our data are consistent with reduced production and secretion of the Q252X variant compared with WT apoA-V after AAV expression in mouse liver.

### Recombinant apoA-V Q252X effectively binds to and exchanges with plasma lipoproteins and lowers plasma TG

We used recombinant proteins to directly compare the functional properties of WT and Q252X apoA-V without the bias of differential expression. ^125^I labeled, lipid-free WT apoA-V protein aggregates upon iodination and is unable to associate with lipoproteins in *apoa5* KO or human plasma **(Figure 5A, 5B, S9A, S9B)**. Surprisingly, ^125^I labeled, lipid-free apoA-V Q252X protein associates with VLDL, LDL and HDL particles when incubated with both plasmas **(Figure 5A, 5B, S9A, S9B)**. We speculated that the absence of the C-terminal hydrophobic region might increase the stability of apoA-V Q252X in monomeric form, limiting protein aggregation in water buffers. To test this hypothesis, we performed a DMPC clearance assay. In buffer-only control incubations without apolipoprotein, DMPC vesicle turbidity remains at baseline **(Figure 5C)**.

**Figure 5.**
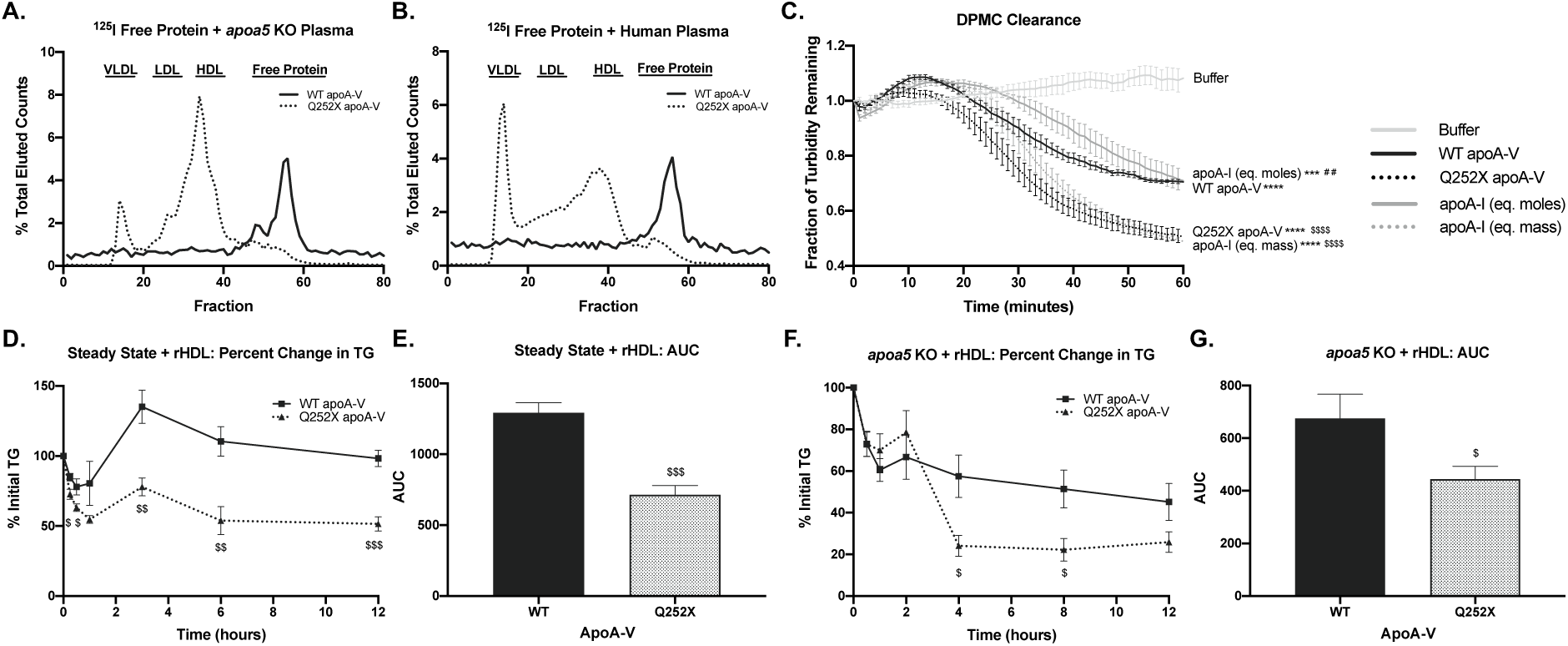
ApoA-V Q252X associates with lipoproteins.^125^I radiolabeled WT or Q252X apoA-V recombinant protein was incubated with **A.** *apoa5* KO and **B.** human plasma then separated by FPLC. Data shown as percent total eluted counts. Fractions corresponding to the elution range of VLDL, LDL and HDL particles are indicated. **C.** DMPC clearance for equivalent moles WT apoA-V and apoA-V Q252X recombinant protein (N=5). **D.** *apoa5* KO mice expressing WT or Q252X apoA-V were administered 5μg WT or Q252X apoA-V synthetic HDL (rHDL). Percent initial TG over time. (N=5-6). **E.** Area under the curve (AUC) for mice shown in G. **F.** *apoa5* KO mice were administered 20μg WT or Q252X apoA-V rHDL. Percent initial TG over time (N=7). **G.** AUC for mice shown in F. **A-B.** Points indicate single value. **C-G.** Mean with SEM (brackets). Unpaired 2-tailed t-test. p<0.001 = ***, p<0.0001 = **** vs. Buffer. p<0.05 = $, p<0.01 = $$, p<0.001 = $$$, p<0.0001 = $$$$ vs. WT. p<0.01 = ## vs. Q252X. apoA-I, apolipoprotein A-I; apoA-V, apolipoprotein A5; auc, area under the curve; DMPC, dimyristoyl phosphatidylcholine; FPLC, fast protein liquid chromatography; HDL, high-density lipoprotein; KO, knockout; LDL, low- density lipoprotein; rHDL, synthetic HDL with recombinant apoA-V protein; TG, triglyceride; VLDL, very low-density lipoprotein.

Addition of apoA-V Q252X decreases DMPC turbidity to a greater extent than equivalent moles of WT apoA-V or apoA-I **(Figure 5C)**. These data suggest that apoA-V Q252X is more readily exchangeable and is associated with more rapid remodeling of DMPC vesicles.

Since our HX MS structures indicated that WT apoA-V becomes more helical and more stable when associated with lipid, we made synthetic HDL containing recombinant human WT or Q252X apoA-V protein (rHDL). Both WT and Q252X apoA-V rHDL preparations fully incorporated the available recombinant protein and made similar, lipoprotein-sized particles **(Figure S10)**. We delivered these rHDL particles to *apoa5 KO* mice expressing WT or Q252X apoA-V (via AAV) and measured plasma TG over time to determine whether the truncation mutant can enhance LPL TG hydrolysis **(Figure S11A)**. WT apoA-V-expressing mice that received 5μg WT apoA-V rHDL show a rapid decrease in plasma TG over one hour **(Figure 5D)**. This decrease is followed by a compensatory increase in plasma TG through three hours and a final decline to 100% of initial plasma TG after 12 hours. ApoA-V Q252X-expressing mice that received equivalent moles apoA-V Q252X rHDL show a rapid and greater decrease in plasma TG over the first hour when compared to WT apoA-V **(Figure 5D)**. Moreover, the compensatory increase in plasma TG through three hours is smaller than observed with WT apoA-V rHDL. 12 hours after injection, mice that received the variant rHDL plateau at 50% of initial plasma TG **(Figure 5D)**. This larger percent plasma TG decrease is highlighted by the significantly reduced area under curve (AUC) observed in apoA-V Q252X mice **(Figure 5E)**.

We also performed a complementary experiment to assess the effect of apoA-V Q252X on LPL TG hydrolysis *in vivo* without the confounder of pre-existing expression **(Figure S11B)**. *Apoa5* KO mice that received 20μg WT apoA-V rHDL show a rapid decrease in plasma TG during the first hour, followed by a compensatory increase in plasma TG **(Figure 5F)**. Through 12 hours, mice which received WT rHDL plateau to 50% initial plasma TG. *apoa5* KO mice that received equivalent moles apoA-V Q252X rHDL also show a rapid initial decrease then spike in plasma TG **(Figure 5F)**. Four hours post-injection, plasma TG in these apoA-V Q252X rHDL-injected mice plummet to 30% of their initial concentrations, where they remain through 12 hours. This robust decline in plasma TG is reflected in the reduced AUC for apoA-V Q252X rHDL **(Figure 5G)**.

## Discussion

We used HX MS to define the first experimentally-derived secondary structure of lipid-free and lipid-associated WT apoA-V. ApoA-V contains multiple helical segments that allow for a highly dynamic and flexible structure. When apoA-V interacts with lipids, its helical content increases. This favorable free energy change likely drives the binding of the exchangeable apoA-V to lipid-water interfaces like TRL and HDL particles. Additionally, apoA-V helix Pf values increase markedly upon interaction with lipids, indicating that apoA-V structure is stabilized when lipid-bound. The *α*-helix spanning residues 311-331 is highly amphipathic and forms a hydrophobic face at the C-terminus. Moreover, AlphaFold modeling of WT apoA-V largely agrees with our highly helical structure and supports the presence of a hydrophobic region at the C-terminus.

To determine the relevance of this region in humans, we examined the lipid and apolipoprotein profiles of human apoA-V Q252X variant carriers (rs149808404), where truncation is predicted to remove the entire C-terminal hydrophobic face. We identified this variant in the PMBB, a large biobank of genomic data linked to Electronic Health Record phenotype data. Carriers of apoA-V Q252X have elevated plasma TG and elevated TG to apoB ratio when compared to matched non-carrier controls, which is consistent with LoF and supportive of apoA-V’s role in lipolysis via mechanisms including inhibition of ANGPTL3/8 inhibition of LPL activity^25, 26^. Carriers of apoA-V Q252X in two other biobanks also have elevated plasma TG.

To examine the phenotypic effects of this variant in isolation, we used AAV vectors to express WT or Q252X apoA-V in *apoa5* KO mice. Mice expressing the truncation variant have elevated plasma TG levels when compared to mice expressing WT apoA-V, recapitulating the LoF phenotype observed in human variant carriers. Liver mRNA abundance and plasma protein were substantially reduced, suggesting an effect on production and secretion of the Q252X variant.

Next, we functionally compared WT and Q252X apoA-V side-by-side using recombinant protein. WT apoA-V is difficult to solubilize, and methods to prepare functional recombinant apoA-V at high concentrations and physiological pH were only recently described by Castleberry et al.^33^. In our hands, lipid-free WT apoA-V recombinant protein could not de-aggregate after iodination and was unable to associate with lipoproteins. Surprisingly, however, lipid-free recombinant Q252X apoA-V protein not only associated with lipoproteins, but also decreased DMPC turbidity better than WT apoA-V. It is known that the dissociation of oligomeric exchangeable apolipoproteins into the monomeric state enhances the rate of the association with DMPC vesicles^69^. Our data suggest that removal of the apoA-V C-terminal hydrophobic face limits the formation of multimeric protein aggregates and stabilizes Q252X apoA-V in monomeric form. This ultimately increases the stability of Q252X apoA-V in water buffers and facilitates its exchange between circulating lipoproteins.

Considering our HX MS data, where WT apoA-V is more helical and more stable when associated with lipid, we made rHDL particles with either WT or Q252X apoA-V. WT apoA-V readily formed lipoprotein-sized particles. Q252X apoA-V also made similar lipoprotein-sized particles, suggesting that the central helix bundle^41^, which is unaffected by the truncation, provides sufficient hydrophobic interactions. *In vivo*, injection of either WT or Q252X rHDLs rapidly decreased plasma TG. This activity suggests that the C-terminal region (aa 252-343) is not necessary for apoA-V intravascular lipolytic activity and that aa 1-251 may retain the as-of-yet undefined domain responsible for interaction with ANGPTL3/8 and promoting LPL activity.

ApoA-V Q252X likely produces a LoF phenotype when endogenously expressed due to reduced mRNA expression (and possibly reduced protein secretion) leading to reduced apoA-V Q252X mass in plasma. The premature stop codon in the Q252X mRNA may destabilize the transcript at translation and decrease steady-state abundance^67, 68^. WT apoA-V protein already circulates at a remarkably low concentration in human plasma (∼150 ng/ml)^15^. Thus, while the apoA-V C-terminus does not appear essential for its LPL-promoting activity in circulation, this hydrophobic region may be important for intracellular lipid binding and proper expression and secretion of the protein.

Others have described engineered and naturally-occurring truncation variants which are associated with hypertriglyceridemia in humans^70–73^. Beckstead et al. examined a truncated apoA-V with amino acids 1-292^40^. This more distal truncation decreased DMPC clearance two-fold, though not completely. Wong-Mauldin et al. studied a more severely truncated apoA-V with amino acids 1-146^70^. Interestingly, this variant forms lipoprotein particles, solubilizes DMPC, and associates with plasma lipoproteins, though to a lesser extent than WT apoA-V. Like apoA-V Q252X, apoA-V 1-146 may be more soluble (monomeric).

We describe the opposite phenotypic and functional consequences for the premature truncation of apoA-V at residue 252. Several studies confirm apoA-V’s role in determining plasma TG levels^16–18^ and risk of hyperTG-induced acute pancreatitis^19–21^ and coronary artery disease^22–24^. Moreover, recent work describes apoA-V’s inhibitory role in ANGPTL3/8-mediated inhibition of LPL TG hydrolysis^25, 26^. Functional analysis of apoA-V Q252X, a rare, naturally occurring variant, has provided valuable insight into apoA-V structure-function and protein biochemistry. Evaluation of this and other variants can be leveraged for the development of novel apoA-V-centered plasma TG-lowering therapeutics.

## Data Availability

Human genetic association data will be made publicly available and experimental data will be made available upon request.

## Acknowledgments

We thank the patients who consented to the inclusion of their samples and clinical data in the PMBB for research. We thank the University of Pennsylvania Vector Core for providing gene vectors used in this study. We gratefully acknowledge Marjorie Risman for identifying matched, non-carrier controls in the PMBB, Kate Townsend Creasy, PhD for her expertise in isolating nuclei from frozen liver, and John Millar, PhD for iodinating recombinant proteins and rHDL. We also acknowledge Linda Morell, Aisha Wilson, Edwige Edouard, Maosen Sun, Debra Cromley, and Amrith Rodrigues for technical assistance, and Jeff Billheimer, PhD, Donna Conlon, PhD, Carolin V. Schneider, MD and Sissel Lund-Katz, PhD for helpful discussion.

## Author Contributions

### Sources of Funding

This work was supported by National Institutes of Health grants R01 HL133502 (to DJR) and National Institutes of Health Fellowship F31 HL149162 (to SS). MVP research is based on data from the Million Veteran Program, Office of Research and Development, Veterans Health Administration, and was supported by award #I01- BX003362 (MV) and the Corporal Michael J. Crescenz VA Medical Center. This publication does not represent the views of the Department of Veteran Affairs or the United States Government. MGL is supported by the Institute for Translational Medicine and Therapeutics of the Perelman School of Medicine at the University of Pennsylvania, the NIH/NHLBI National Research Service Award postdoctoral fellowship (T32HL007843), and the Measey Foundation.

### Disclosures

DJR is a consultant for Alnylam, Novartis, Pfizer, and Verve.

## Supplemental Material

Tables S1-S7

Figures S1-S11

## Abbreviations

AAV: Adeno-associated virus
APOA1: Apolipoprotein A1 (protein, apoA-I)
APOA5: Apolipoprotein A5 (protein, apoA-V)
APOB: Apolipoprotein B (protein, apoB)
APOC3: Apolipoprotein C3 (protein, apoC-III)
ANGPTL3/8: Angiopoietin-like-protein 3/8
AUC: Area under curve
DMPC: Dimyristoyl phosphatidylcholine
GLGC: Global Lipids Genetics Consortium
GPIHBP1: Glycosylphosphatidylinositol anchored high density lipoprotein binding protein 1
HDL-C: High-density lipoprotein cholesterol
HyperTG: Hypertriglyceridemia
HX MS: Hydrogen-deuterium exchange mass spectrometry
KO: Knockout
LoF: Loss-of-function
MVP: Million Veteran Program
Pf: Protection factor
PMBB: Penn Medicine Biobank
POPC: Palmitoyl-oleoyl phosphatidylcholine
TG: Triglyceride
TRL: Triglyceride-rich lipoprotein
UKBB: UK Biobank
VLDL: Very low-density lipoprotein
WT: Wildtype

## Supplemental Materials

### Tables

**Table S1.**
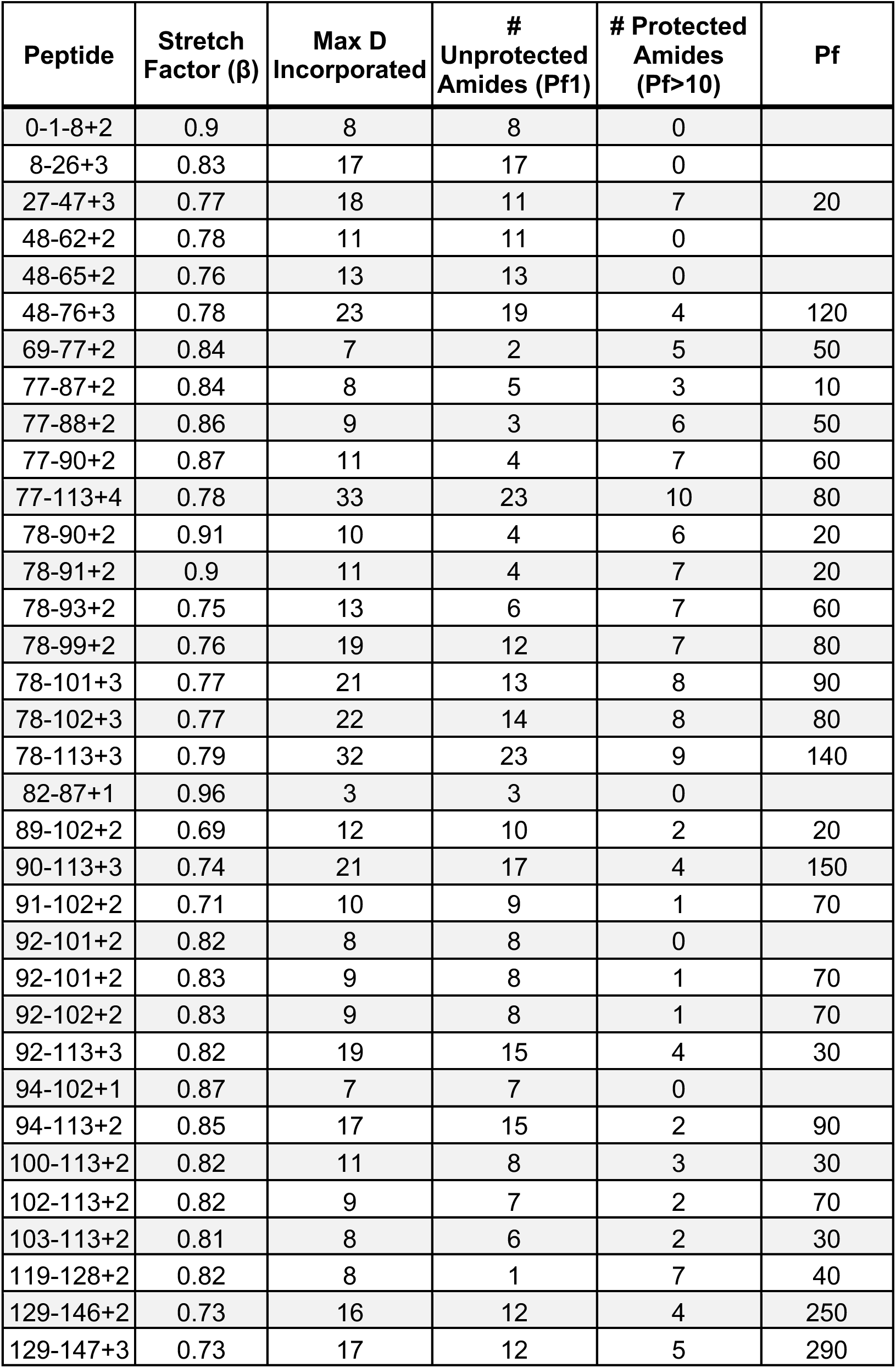
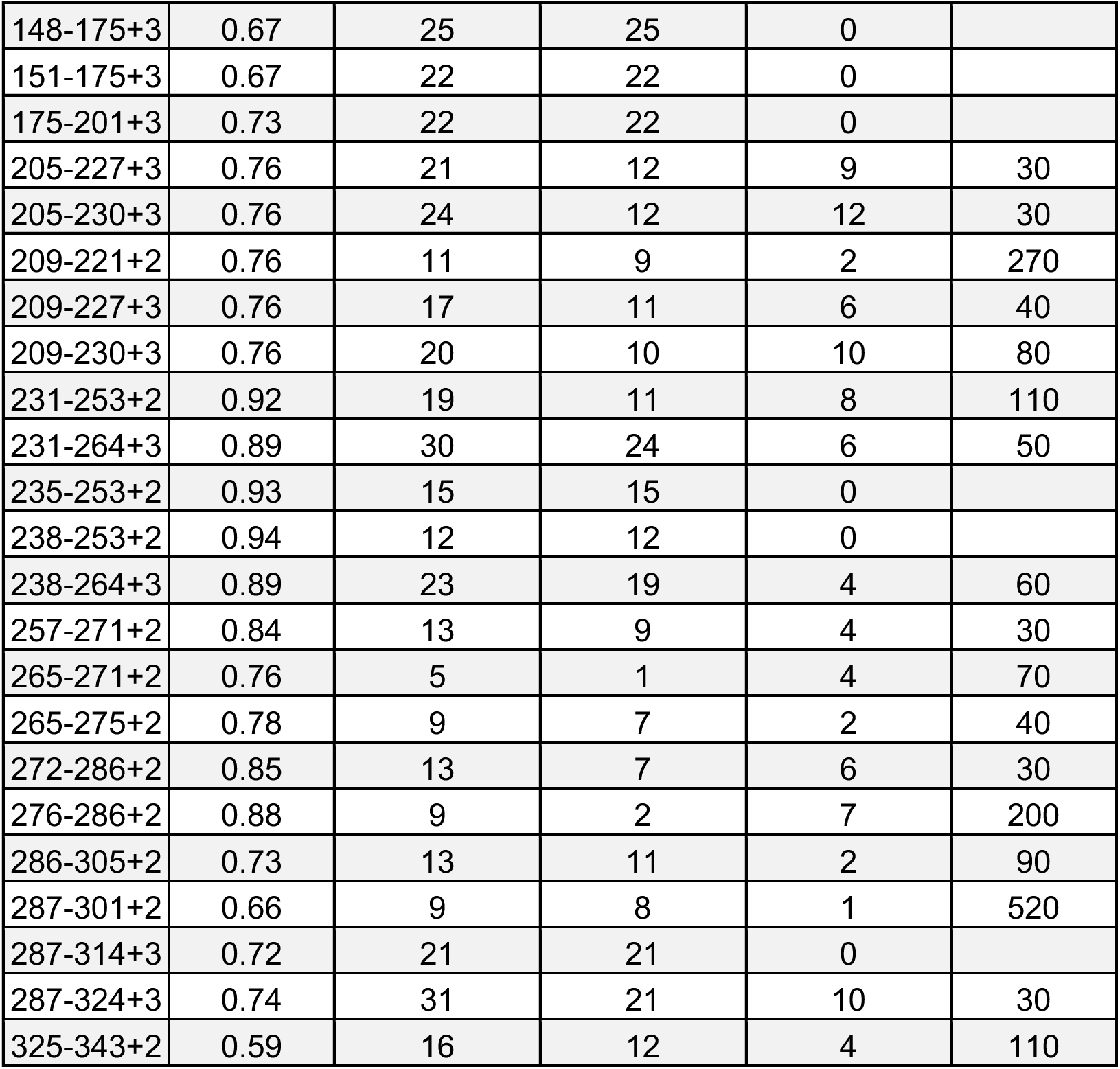
HX kinetics of lipid-free apoA-V peptides. The HX time-courses obtained from the mass spectra of the 56 peptides included in Figure S1 were fitted with one or two exponentials to derive the tabulated protection factors (Pf) and numbers of amides involved in each kinetic phase. The fitting to the exponential equations was performed with IGOR Pro (Wavemetrics Inc.). apoA-V, apolipoprotein A5; HX MS, hydrogen-deuterium exchange mass spectrometry; Pf, protection factor.

| Peptide | Stretch Factor ($\beta$) | Max D Incorporated | # Unprotected Amides (Pf1) | # Protected Amides (Pf>10) | Pf |
| --- | --- | --- | --- | --- | --- |
| 0-1-8+2 | 0.9 | 8 | 8 | 0 |  |
| 8-26+3 | 0.83 | 17 | 17 | 0 |  |
| 27-47+3 | 0.77 | 18 | 11 | 7 | 20 |
| 48-62+2 | 0.78 | 11 | 11 | 0 |  |
| 48-65+2 | 0.76 | 13 | 13 | 0 |  |
| 48-76+3 | 0.78 | 23 | 19 | 4 | 120 |
| 69-77+2 | 0.84 | 7 | 2 | 5 | 50 |
| 77-87+2 | 0.84 | 8 | 5 | 3 | 10 |
| 77-88+2 | 0.86 | 9 | 3 | 6 | 50 |
| 77-90+2 | 0.87 | 11 | 4 | 7 | 60 |
| 77-113+4 | 0.78 | 33 | 23 | 10 | 80 |
| 78-90+2 | 0.91 | 10 | 4 | 6 | 20 |
| 78-91+2 | 0.9 | 11 | 4 | 7 | 20 |
| 78-93+2 | 0.75 | 13 | 6 | 7 | 60 |
| 78-99+2 | 0.76 | 19 | 12 | 7 | 80 |
| 78-101+3 | 0.77 | 21 | 13 | 8 | 90 |
| 78-102+3 | 0.77 | 22 | 14 | 8 | 80 |
| 78-113+3 | 0.79 | 32 | 23 | 9 | 140 |
| 82-87+1 | 0.96 | 3 | 3 | 0 |  |
| 89-102+2 | 0.69 | 12 | 10 | 2 | 20 |
| 90-113+3 | 0.74 | 21 | 17 | 4 | 150 |
| 91-102+2 | 0.71 | 10 | 9 | 1 | 70 |
| 92-101+2 | 0.82 | 8 | 8 | 0 |  |
| 92-101+2 | 0.83 | 9 | 8 | 1 | 70 |
| 92-102+2 | 0.83 | 9 | 8 | 1 | 70 |
| 92-113+3 | 0.82 | 19 | 15 | 4 | 30 |
| 94-102+1 | 0.87 | 7 | 7 | 0 |  |
| 94-113+2 | 0.85 | 17 | 15 | 2 | 90 |
| 100-113+2 | 0.82 | 11 | 8 | 3 | 30 |
| 102-113+2 | 0.82 | 9 | 7 | 2 | 70 |
| 103-113+2 | 0.81 | 8 | 6 | 2 | 30 |
| 119-128+2 | 0.82 | 8 | 1 | 7 | 40 |
| 129-146+2 | 0.73 | 16 | 12 | 4 | 250 |
| 129-147+3 | 0.73 | 17 | 12 | 5 | 290 |
| 148-175+3 | 0.67 | 25 | 25 | 0 |  |
| 151-175+3 | 0.67 | 22 | 22 | 0 |  |
| 175-201+3 | 0.73 | 22 | 22 | 0 |  |
| 205-227+3 | 0.76 | 21 | 12 | 9 | 30 |
| 205-230+3 | 0.76 | 24 | 12 | 12 | 30 |
| 209-221+2 | 0.76 | 11 | 9 | 2 | 270 |
| 209-227+3 | 0.76 | 17 | 11 | 6 | 40 |
| 209-230+3 | 0.76 | 20 | 10 | 10 | 80 |
| 231-253+2 | 0.92 | 19 | 11 | 8 | 110 |
| 231-264+3 | 0.89 | 30 | 24 | 6 | 50 |
| 235-253+2 | 0.93 | 15 | 15 | 0 |  |
| 238-253+2 | 0.94 | 12 | 12 | 0 |  |
| 238-264+3 | 0.89 | 23 | 19 | 4 | 60 |
| 257-271+2 | 0.84 | 13 | 9 | 4 | 30 |
| 265-271+2 | 0.76 | 5 | 1 | 4 | 70 |
| 265-275+2 | 0.78 | 9 | 7 | 2 | 40 |
| 272-286+2 | 0.85 | 13 | 7 | 6 | 30 |
| 276-286+2 | 0.88 | 9 | 2 | 7 | 200 |
| 286-305+2 | 0.73 | 13 | 11 | 2 | 90 |
| 287-301+2 | 0.66 | 9 | 8 | 1 | 520 |
| 287-314+3 | 0.72 | 21 | 21 | 0 |  |
| 287-324+3 | 0.74 | 31 | 21 | 10 | 30 |
| 325-343+2 | 0.59 | 16 | 12 | 4 | 110 |

**Table S2.** HX kinetics of lipid-associated apoA-V peptides (DMPC vesicles, 10/1 w/w DMPC/apoA-V). The HX time-courses obtained from the mass spectra of the 70 peptides included in Figure S2 were fitted with one or two exponentials to derive the tabulated protection factors (Pf) and numbers of amides involved in each kinetic phase. The fitting to the exponential equations was performed with IGOR Pro (Wavemetrics Inc.). apoA-V, apolipoprotein A5; DMPC, dimyristoyl phosphatidylcholine; HX MS, hydrogen-deuterium exchange mass spectrometry; Pf, protection factor.

| Peptide | Stretch Factor ($\beta$) | Max D Incorporated | # Unprotected Amides (Pf1) | # Protected Amides (Pf>10) | Pf |
| --- | --- | --- | --- | --- | --- |
| 0-1-23+3 | 0.83 | 23 | 23 | 0 |  |
| 9-33+3 | 0.82 | 22 | 22 | 0 |  |
| 27-47+3 | 0.77 | 18 | 9 | 9 | 70 |
| 34-47+2 | 0.79 | 12 | 5 | 7 | 60 |
| 38-47+2 | 0.82 | 8 | 3 | 5 | 40 |
| 48-62+2 | 0.78 | 11 | 9 | 2 | 10 |
| 48-65+3 | 0.76 | 13 | 13 | 0 |  |
| 48-68+3 | 0.73 | 15 | 15 | 0 |  |
| 48-69+3 | 0.74 | 16 | 16 | 0 |  |
| 48-76+3 | 0.78 | 23 | 19 | 4 | 80 |
| 78-91+2 | 0.9 | 11 | 4 | 7 | 130 |
| 78-113+4 | 0.79 | 32 | 15 | 17 | 90 |
| 94-102+1 | 0.87 | 7 | 5 | 2 | 90 |
| 100-113+2 | 0.82 | 11 | 5 | 6 | 60 |
| 102-113+2 | 0.82 | 9 | 4 | 5 | 120 |
| 103-113+2 | 0.81 | 8 | 3 | 5 | 80 |
| 114-119+1 | 0.78 | 4 | 4 | 0 |  |
| 117-128+2 | 0.82 | 10 | 5 | 5 | 100 |
| 119-128+2 | 0.82 | 8 | 1 | 7 | 170 |
| 120-128+2 | 0.85 | 7 | 1 | 6 | 230 |
| 129-135+1 | 0.64 | 5 | 2 | 3 | 140 |
| 129-139+2 | 0.75 | 9 | 4 | 5 | 250 |
| 129-146+2 | 0.73 | 16 | 6 | 10 | 190 |
| 147-150+1 | 0.98 | 2 | 2 | 0 |  |
| 147-164+2 | 0.62 | 16 | 8 | 8 | 60 |
| 148-164+3 | 0.63 | 15 | 8 | 7 | 80 |
| 148-167+3 | 0.67 | 18 | 9 | 9 | 20 |
| 148-175+3 | 0.67 | 25 | 18 | 7 | 80 |
| 150-164+3 | 0.64 | 13 | 13 | 0 |  |
| 151-175+3 | 0.67 | 22 | 17 | 5 | 60 |
| 156-175+3 | 0.65 | 17 | 15 | 2 | 10 |
| 165-174+2 | 0.68 | 7 | 7 | 0 |  |
| 165-178+2 | 0.69 | 11 | 11 | 0 |  |
| 165-186+3 | 0.71 | 19 | 19 | 0 |  |
| 168-175+2 | 0.76 | 5 | 5 | 0 | 40 |
| 168-186+3 | 0.73 | 16 | 16 | 0 |  |
| 175-201+3 | 0.73 | 22 | 22 | 0 |  |
| 176-201+3 | 0.72 | 21 | 21 | 0 |  |
| 182-201+3 | 0.73 | 15 | 15 | 0 |  |
| 187-201+3 | 0.74 | 10 | 10 | 0 |  |
| 205-227+3 | 0.76 | 21 | 5 | 16 | 90 |
| 205-230+3 | 0.76 | 24 | 5 | 19 | 100 |
| 209-227+3 | 0.76 | 17 | 6 | 11 | 90 |
| 209-229+3 | 0.77 | 19 | 5 | 14 | 90 |
| 209-230+3 | 0.76 | 20 | 5 | 15 | 100 |
| 231-253+2 | 0.92 | 19 | 15 | 4 | 80 |
| 231-264+3 | 0.89 | 30 | 19 | 11 | 80 |
| 235-253+2 | 0.93 | 15 | 11 | 4 | 120 |
| 238-253+2 | 0.94 | 12 | 9 | 3 | 120 |
| 238-264+3 | 0.89 | 23 | 15 | 8 | 150 |
| 257-271+2 | 0.84 | 13 | 5 | 8 | 80 |
| 265-271+2 | 0.76 | 5 | 0 | 5 | 170 |
| 265-275+2 | 0.78 | 9 | 4 | 5 | 70 |
| 272-286+2 | 0.85 | 13 | 5 | 8 | 170 |
| 276-283+1 | 0.84 | 6 | 6 | 0 |  |
| 276-286+2 | 0.88 | 9 | 3 | 6 | 290 |
| 286-305+2 | 0.73 | 13 | 10 | 3 | 50 |
| 287-301+2 | 0.66 | 9 | 7 | 2 | 50 |
| 287-314+3 | 0.72 | 21 | 21 | 0 |  |
| 287-317+3 | 0.73 | 24 | 20 | 4 | 60 |
| 287-321+3 | 0.75 | 28 | 22 | 6 | 80 |
| 287-324+3 | 0.74 | 31 | 19 | 12 | 60 |
| 287-324+4 | 0.74 | 31 | 19 | 12 | 190 |
| 302-321+3 | 0.80 | 18 | 11 | 7 | 90 |
| 302-324+3 | 0.77 | 21 | 12 | 9 | 70 |
| 306-321+2 | 0.75 | 14 | 7 | 7 | 70 |
| 306-321+3 | 0.75 | 14 | 9 | 5 | 160 |
| 325-342+2 | 0.59 | 16 | 5 | 11 | 150 |
| 325-342+3 | 0.59 | 16 | 11 | 5 | 480 |
| 325-343+2 | 0.59 | 16 | 11 | 5 | 280 |

**Table S3.** Characteristics of human APOA5 variant carriers and matched controls (non-carriers). APOA5 variant carriers and age-, sex-, BMI-, ancestry-, and history of diabetes mellitus diagnosis-matched controls (non-carriers) were identified from exome-wide genotyping in the PMBB. Sex (male/female), age (years, mean ± standard deviation), Body Mass Index (BMI, kg/m^2^, mean ± standard deviation), ancestry (Asian/Black/White), and diabetes mellitus status (DM, yes/no). apoa5, apolipoprotein A5; BMI, body mass index; DM, diabetes mellitus.

| Variant |  | G162C | Q252X |
| --- | --- | --- | --- |
| N | Carriers | 73 | 13 |
|  | Non-Carriers | 73 | 39 |
| Male/Female | Carriers | 28/45 | 9/4 |
|  | Non-Carriers | 28/45 | 27/12 |
| Age (years) | Carriers | 53.8±15.9 | 57.8±15.8 |
|  | Non-Carriers | 53.4±14.9 | 60.9±8.0 |
| BMI (kg/m<sup>2</sup>) | Carriers | 31.4±8.4 | 26.8±3.3 |
|  | Non-Carriers | 31.6±8.4 | 27.0±2.7 |
| Ancestry<br>(Asian/Black/White) | Carriers | 8/56/9 | 0/0/13 |
|  | Non-Carriers | 8/56/9 | 0/0/39 |
| DM (yes/no) | Carriers | 24/49 | 2/11 |
|  | Non-Carriers | 24/49 | 6/33 |

**Table S4.** Plasma lipid and apolipoprotein measurements of human *APOA5* variant carriers and matched controls (non-carriers). *APOA5* variant carriers and age-, sex-, BMI-, ancestry-, and history of diabetes mellitus diagnosis-matched controls (non-carriers) were identified from exome-wide genotyping in the Penn Medicine BioBank. Plasma measurements of ApoA-I, apoC-II, apoC-III, TC, LDL-C, Non-HDL-C, HDL-C, FC, HDL-C:ApoA-I ratio, and PL. " (standardized regression coefficient; slope), SE and significance determined by linear regression analysis adjusted by SNP, age, sex, race, BMI and diabetes mellitus status. P < 0.01 = **; p < 0.001 = ***. ApoA-I, apolipoprotein A-I; apoC-II, apolipoprotein C-II; apoC-III, apolipoprotein C-III; TC, total cholesterol; LDL-C, low-density lipoprotein cholesterol; non-HDL-C, non-high-density lipoprotein cholesterol; HDL-C, high density lipoprotein cholesterol; FC, free cholesterol; PL, phospholipid; SE, standard error.

| Variant |  | G162C | Q252X |
| --- | --- | --- | --- |
| ApoA-I | $\beta$ | +2.26 | +3.62 |
|  | SE | 4.55 | 9.11 |
| ApoC-II | $\beta$ | +0.19 | +2.04 ** |
|  | SE | 0.36 | 0.62 |
| ApoC-III | $\beta$ | +1.34 | +5.55 *** |
|  | SE | 0.83 | 1.43 |
| TC | $\beta$ | -2.27 | +17.72 |
|  | SE | 6.63 | 13.80 |
| LDL-C | $\beta$ | -5.21 | +6.78 |
|  | SE | 5.55 | 11.52 |
| Non-HDL-C | $\beta$ | +1.50 | +24.39 |
|  | SE | 6.62 | 13.92 |
| HDL-C | $\beta$ | -3.78 | -6.67 |
|  | SE | 2.19 | 4.62 |
| FC | $\beta$ | -1.91 | +5.98 |
|  | SE | 1.90 | 3.73 |
| HDL-C:ApoA-I | $\beta$ | -0.03 *** | -0.05 *** |
|  | SE | 0.01 | 0.01 |
| PL | $\beta$ | -1.44 | +20.60 |
|  | SE | 6.35 | 12.21 |
| N | Carriers | 73 | 13 |
|  | Non-Carriers | 73 | 39 |

**Table S5.** Human apoA-V G162C and Q252X are nominally associated with elevated plasma TG and decreased HDL-C in MVP. TG and HDL-C estimates were obtained by querying the MVP database for clinically obtained measurements in the Veterans Health Administration electronic health record system. Associations between G162C and Q252X and lipid measurements were performed among participants of each genetic ancestry, accounting for age, sex, and 10 genetic principal components. Linear regression was performed to test associations with lipid levels. Estimate, standard error, p value and samples size provided. AFR, African; apoA-V, apolipoprotein A5; EUR, European; HDL-C, high-density lipoprotein cholesterol; HIS, Hispanic; MVP, Million Veteran Program; TG, triglyceride.

| Lipid | Variant | Ancestry | Estimate | Std. Error | P Value | Sample Size |
| --- | --- | --- | --- | --- | --- | --- |
| TG | G162C | AFR | +33.967 | 3.993 | 1.82E-17 | 101469 |
|  |  | EUR | +23.138 | 7.008 | 9.61E-04 | 368884 |
|  |  | HIS | +28.282 | 14.280 | 4.77E-02 | 43742 |
|  | Q252X | AFR | -13.044 | 93.191 | 8.89E-01 | 101469 |
|  |  | EUR | +89.564 | 11.553 | 9.05E-15 | 368884 |
|  |  | HIS | -35.413 | 84.629 | 6.76E-01 | 43742 |
| HDL-C | G162C | AFR | -2.412 | 0.683 | 4.13E-04 | 102015 |
|  |  | EUR | -3.916 | 0.960 | 4.53E-05 | 372607 |
|  |  | HIS | -2.187 | 1.550 | 1.58E-01 | 43829 |
|  | Q252X | AFR | +8.379 | 15.977 | 6.00E-01 | 102015 |
|  |  | EUR | -5.663 | 1.586 | 3.58E-04 | 372607 |
|  |  | HIS | +3.902 | 9.186 | 6.71E-01 | 43829 |

**Table S6.** Human apoA-V G162C and Q252X are nominally associated with elevated plasma TG and decreased HDL-C and apoA-I in UKBB. TG, apoB, HDL-C and apoA-I estimates were obtained by querying each variant using Variant PheWAS in Genebass. For G162C, alternate allele count = 135, allele number = 735926 and alternate allele frequency = 0.00018. For Q252X, alternate allele count = 72, allele number = 735924, and alternate allele frequency = 0.00010. apoA-I, apolipoprotein A1; apoA-V, apolipoprotein A5; apoB, apolipoprotein B; HDL-C, high-density lipoprotein cholesterol; TG, triglyceride; UKBB, UK Biobank.

| Lipid | Variant | $\beta$ | P Value |
| --- | --- | --- | --- |
| TG | G162C | +0.42182 | 1.18E-07 |
|  | Q252X | +0.98443 | 3.19E-19 |
| ApoB | G162C | +0.09138 | 2.64E-01 |
|  | Q252X | +0.21018 | 6.75E-02 |
| HDL-C | G162C | -0.36565 | 2.21E-06 |
|  | Q252X | -0.59038 | 5.78E-08 |
| ApoA-I | G162C | -0.25724 | 1.12E-03 |
|  | Q252X | -0.26114 | 1.84E-02 |

**Table S7.**
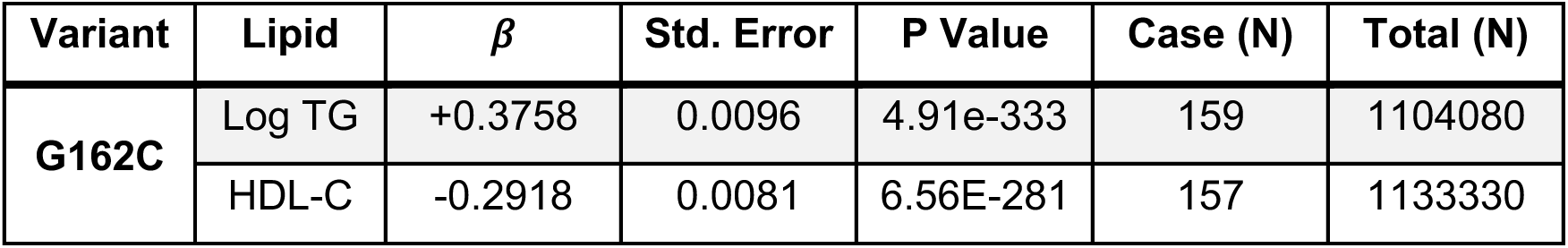
Human apoA-V G162C is associated with elevated plasma TG and decreased HDL-C in GLGC. Log TG and HDL-C estimates were accessed in the Global Lipids Genetics Consortium (GLGC) result files and represent meta-analysis across all ancestries for the entire dataset provided. Pooled alternate allele frequency for log TG = 0.00543 and pooled alternate allele frequency for HDL-C = 0.00752. apoA-V, apolipoprotein A5; GLGC, Global lipids Genetics Consortium; HDL-C, high-density lipoprotein cholesterol; TG, triglyceride.

### Figures

**Figure S1.**
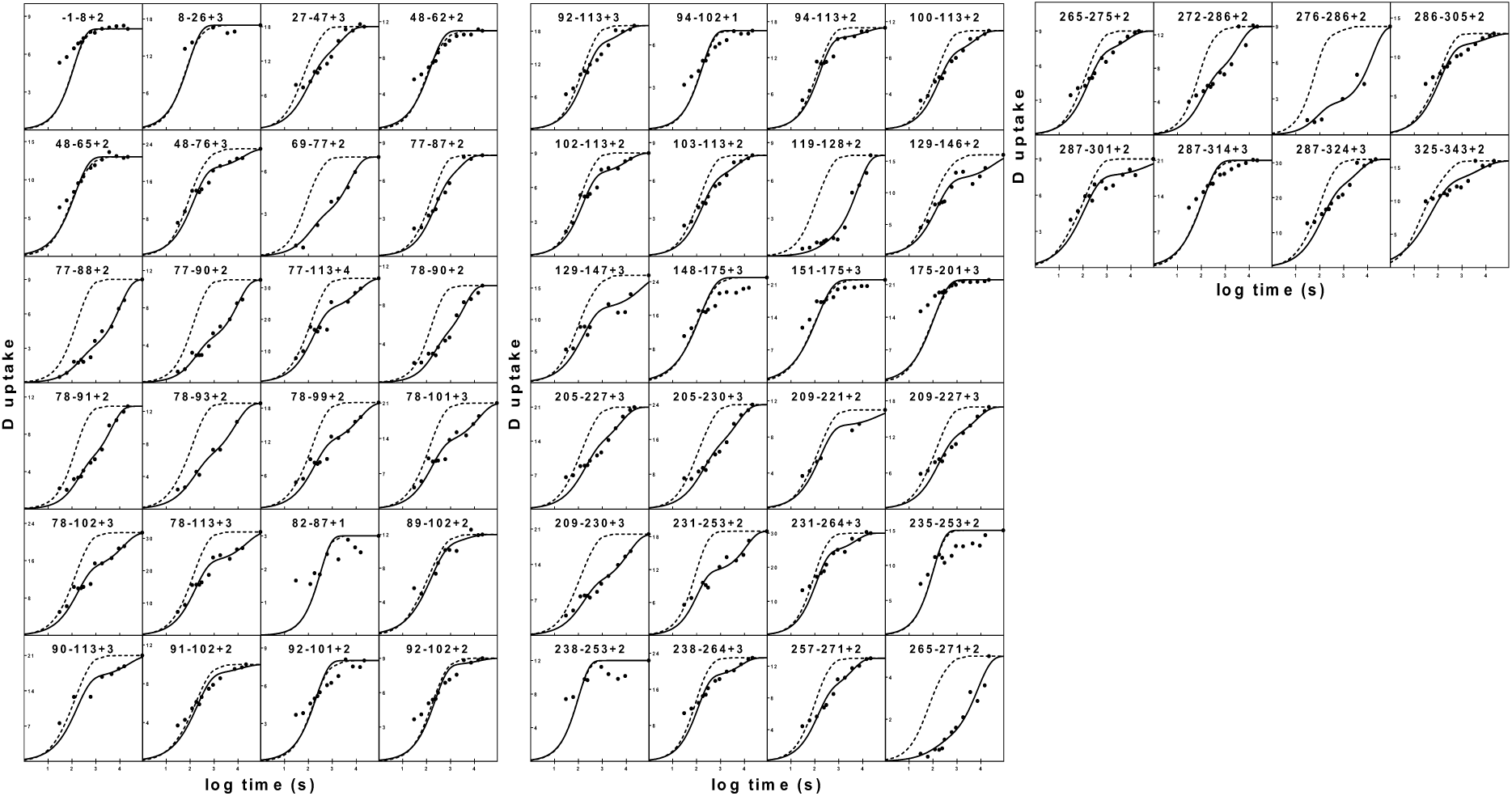
HX kinetic profiles of lipid-free human apoA-V. HX kinetics (pH 3.8, 25°C, 20-40µg/ml) for 56 peptides covering the entire sequence of mature apoA-V. Residue numbers and charge indicated at the top of each panel. Observed HX kinetics of an apoA-V fragment (•) compared to the intrinsic rate for the peptide (dotted line). Time-courses are fitted to either stretched mono-exponential (single cooperatively unfolding segment of secondary structure) or bi-exponential (peptide that spans a helix terminus) rate equations. The kinetic fitting parameters are listed in Table S1. apoA-V, apolipoprotein A5; HX MS, hydrogen-deuterium exchange mass spectrometry.

**Figure S2.**
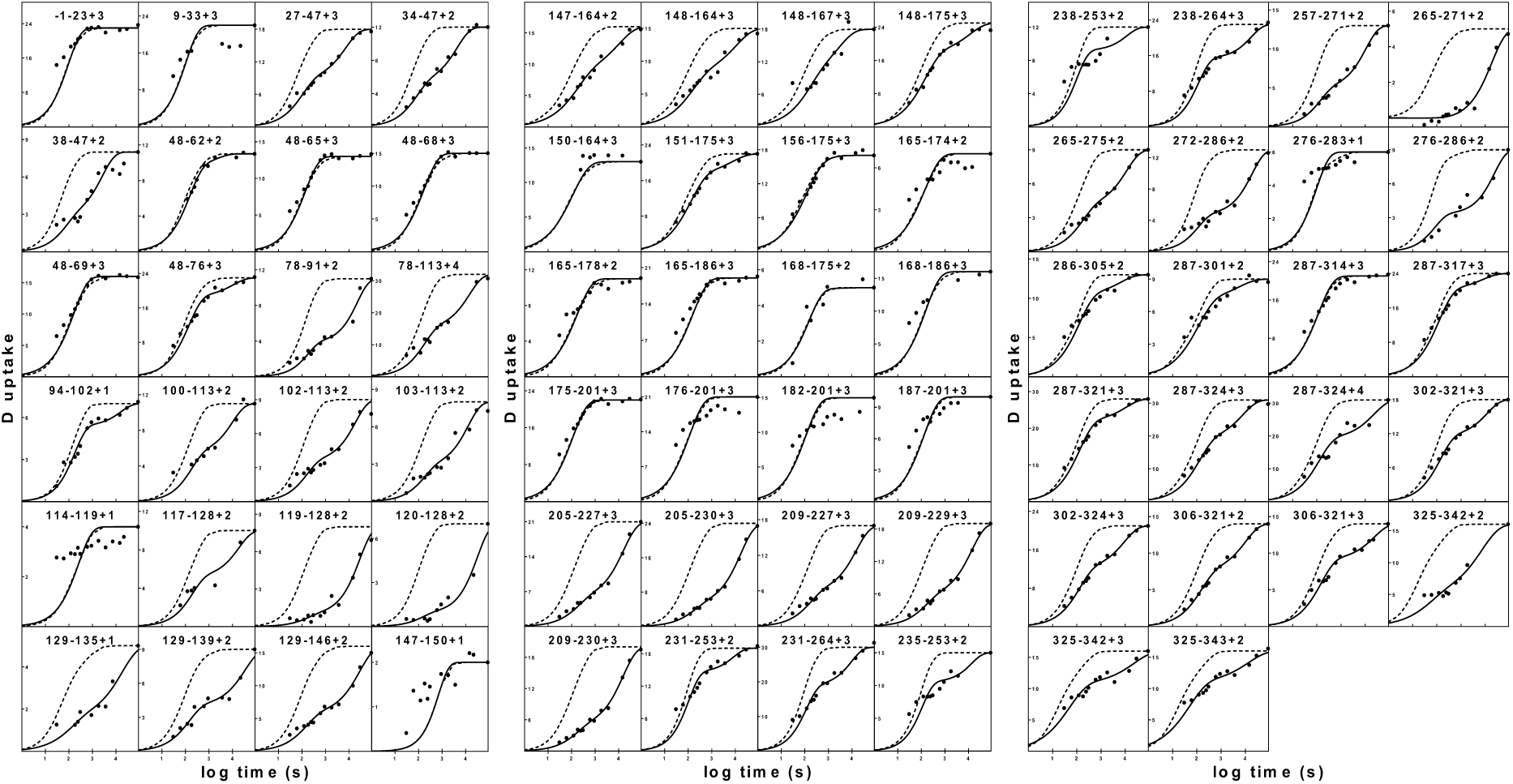
HX kinetic profiles of lipid-associated human apoA-V. HX kinetics (pH 3.8, 25°C, 20- 40µg/ml) for 70 peptides covering the entire sequence of mature apoA-V. Residue numbers and charge indicated at the top of each panel. Observed HX kinetics for apoA-V fragments associated with DMPC vesicles (10/1 w/w DMPC/apoA-V) (•) compared to the intrinsic rate for the peptide (dotted line). Time-courses are fitted to either stretched mono-exponential (single cooperatively unfolding segment of secondary structure) or bi-exponential (peptide that spans a helix terminus) rate equations. The kinetic fitting parameters are listed in Table S2. apoA-V, apolipoprotein A5; DMPC, dimyristoyl phosphatidylcholine; HX MS, hydrogen-deuterium exchange mass spectrometry.

**Figure S3.**
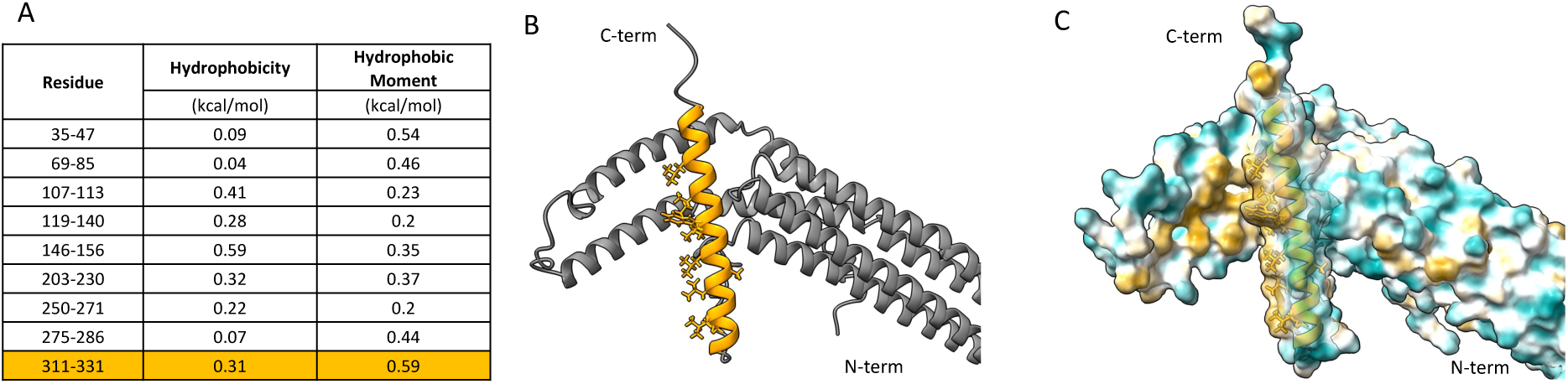
Hydrophobicities and amphiphilicities of α-helices in lipid-associated apoA-V and structure prediction of hydrophobic face. **A.** Table of hydrophobicities and amphiphilicities of α- helices in lipid-associated apoA-V. Residue numbering refers to the helix locations in lipid-bound apoA-V, as determined by HX MS. The C-terminal hydrophobic face, spanning aa 311-331 is highlighted. Values for hydrophobicity and hydrophobic moment were calculated with Heliquest. **A.** Detail of the most C-terminal helix of WT apoA-V. The hydrophobic region (helix encompassing aa 310-335) is highlighted in orange. The side chains of hydrophobic amino acids are shown. **C.** Molecular surface maps of the lipophilic/hydrophilic properties of the amino acids shown in B. Surfaces are color-coded based on Molecular Lipophilicity Potential (MLP), with green being the least lipophilic and gold being the most lipophilic. MLP maps were generated with ChimeraX-1.4. apoA-V, apolipoprotein A5; HX MS, hydrogen-deuterium exchange mass spectrometry.

**Figure S4.**
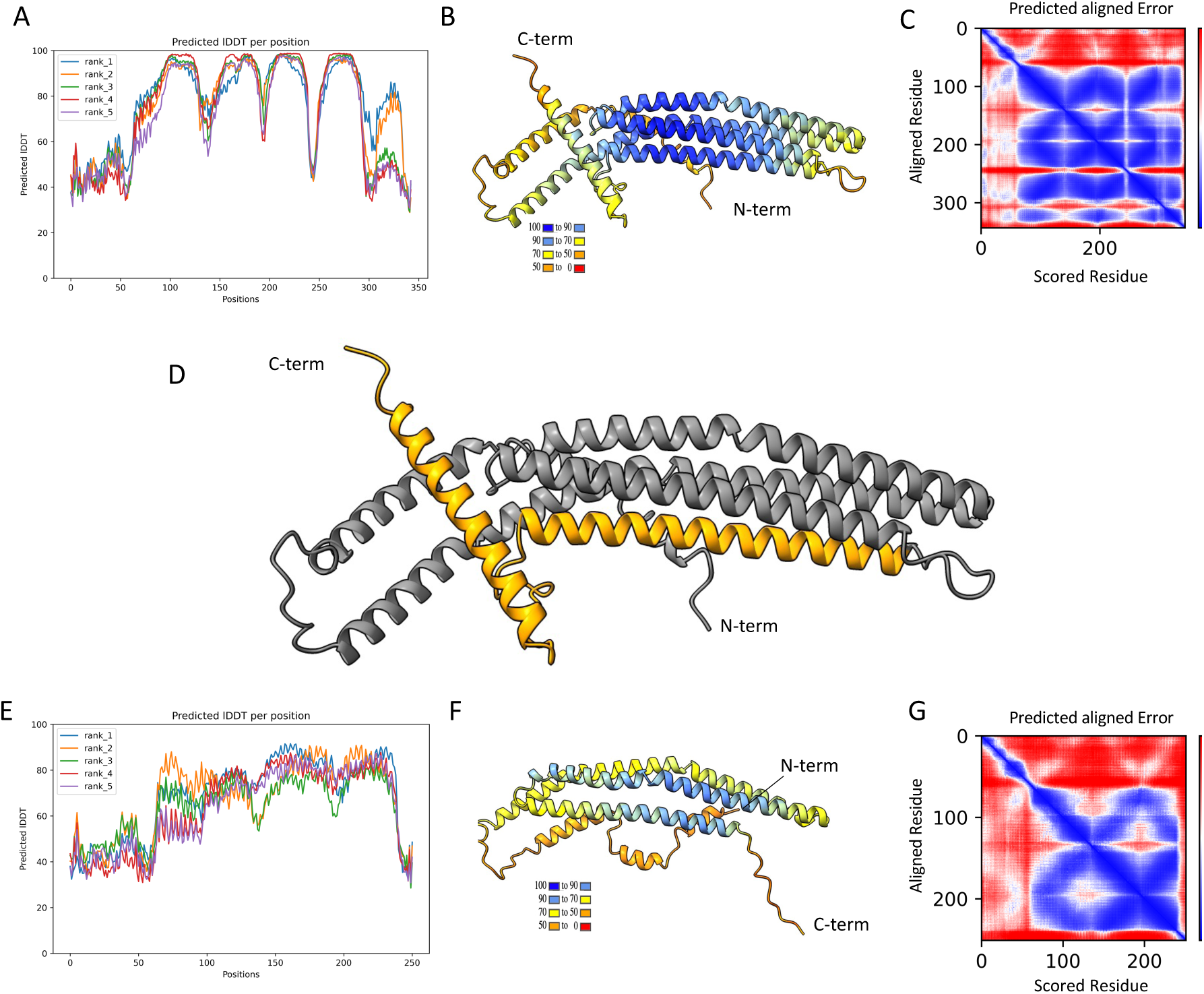
AlphaFold structure predictions of WT and Q252X apoA-V. **A.** Plots of local distance difference test (lDDT) scores (y axis) by residue positions (x axis) for 5 simulated models of WT apoA-V. lDDT scores are a measure of the model confidence^74^. pLDDT > 90 indicates high accuracy. pLDDT between 70 and 90 indicates good accuracy of the backbone prediction. pLDDT between 50 and 70 indicates low confidence. pLDDT between 50 and 0 indicates that the region should not be interpreted and may be disordered. The highest confidence model (rank 1) was selected for further analyses. **B.** Highest confidence model of WT ApoA-V structure, color coded by lDDT score. The key indicates the color codes for different lDDT scores. **C.** Predicted aligned error (PAE) plot for the highest confidence model of WT apoA-V. The PAE is a measure of the model confidence for the relative position of domains. The plot indicates the estimated position error at residue x if the predicted and true structures were aligned on residue y. High PAE for a pair of residues located in two different domains indicates high degree of uncertainty on the relative position of the two domains. **D.** WT apoA-V structure, showing the amino acid sequence (orange) removed upon the premature termination of translation at position 252. **E-G.** AlphaFold structure predictions of Q252X apoA-V. **E.** Plot of local distance difference test (lDDT) scores by residue positions for Q252X apoA-V. **G.** Highest confidence model of Q252X apoA-V structure, color coded by lDDT score. **H.** Predicted aligned error plot for the highest confidence model of Q252X apoA-V. Plots shown in E-G were generated as described for panel A-C. apoA-V, apolipoprotein A5; HX MS, hydrogen-deuterium exchange mass spectrometry; lDDT, local distance difference test; PAE, predicted aligned error.

**Figure S5.**
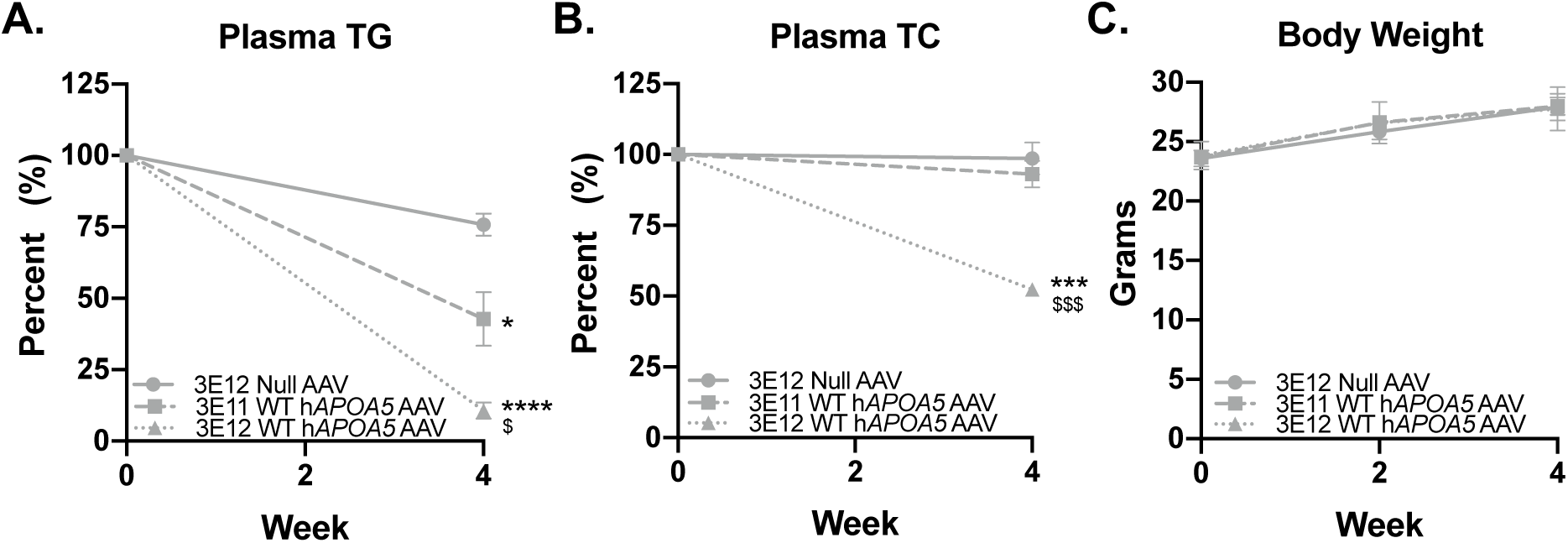
*Apoa5* KO mice exhibit dose dependent decreases in plasma TG after WT *APOA5* AAV injection. *Apoa5* KO mice were fasted 4 hours, bled, then injected with 3E12 Null AAV or 3E11 or 3E12 WT h*APOA5* AAV. **A.** Plasma TG and **B.** Plasma TC at baseline and 4 weeks after AAV administration **C.** Body weight at indicated time-points (N=3-5). Mean with SEM (brackets). Unpaired 2-tailed t-test. p<0.05 = *, p<0.001 = ***, p<0.0001 = **** vs. 3E12 Null. p<0.05 = $, p<0.001 = $$$, = vs. 3E11 WT. apoa5, apolipoprotein A5; KO, knockout; TC, total cholesterol; TG, triglyceride.

**Figure S6.**
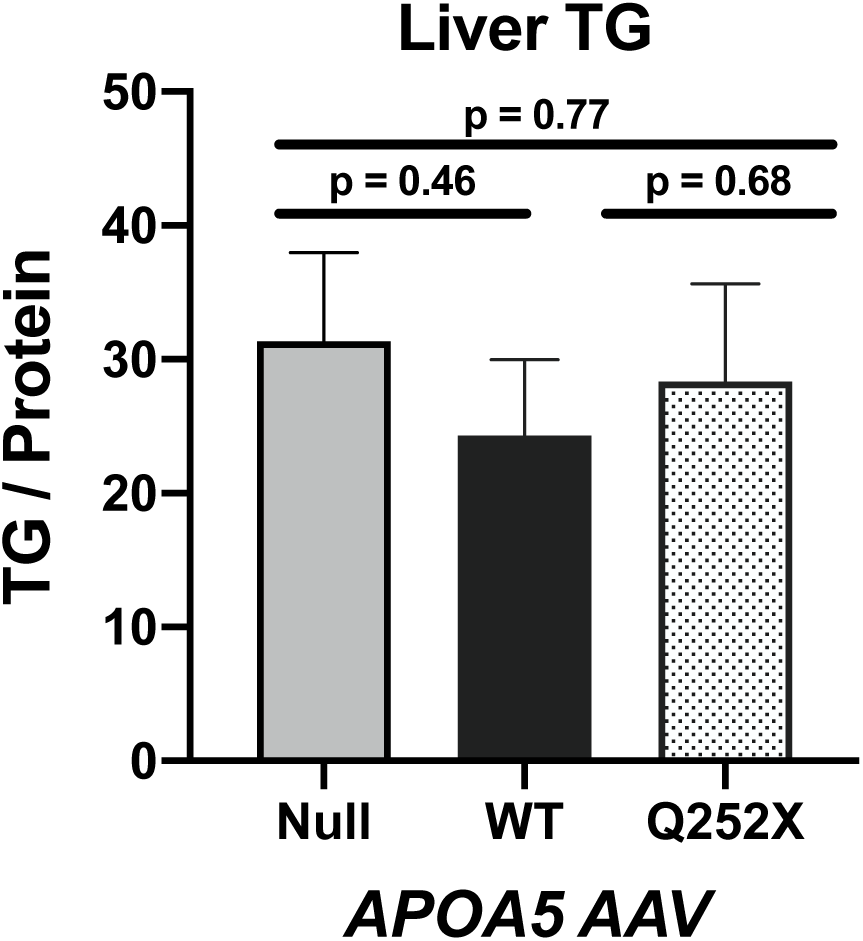
*Apoa5* KO mice administered WT or Q252X h*APOA5* AAV exhibit no changes in liver TG content after four weeks. *Apoa5* KO mice were fasted 4 hours, bled, then injected with 3E11 Null AAV or WT or Q252X h*APOA5* AAV. Liver TG four weeks after AAV injection (N=4-5). Mean with SEM. Unpaired 2-tailed t-test. Apoa5, apolipoprotein A5; KO, knockout; TG, triglyceride.

**Figure S7.**
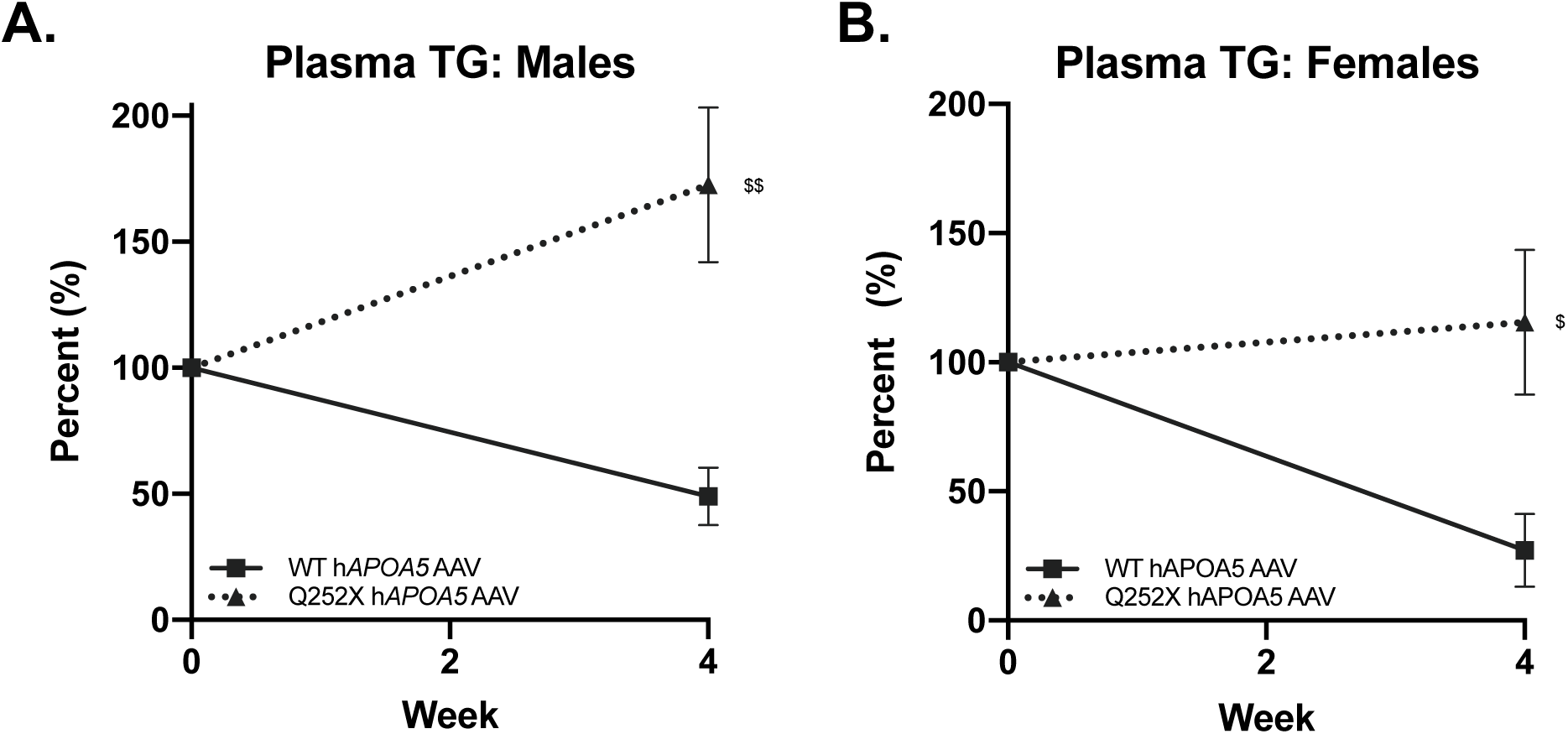
*apoa5* KO mice expressing apoA-V Q252X have higher plasma TG. *apoa5* KO mice were fasted 4 hours, bled, then injected with 3E12 WT or Q252X h*APOA5* AAV. **A.** Male mice plasma TG (N=5-6). **B.** Female mice plasma TG (N=4). Results are expressed as percent of baseline levels. Mean with SEM. Unpaired 2-tailed t-test. p<0.05 = $, p<0.01 = $$ vs. WT. apoA-V, apolipoprotein A5; KO, knockout; TG, triglyceride.

**Figure S8.**
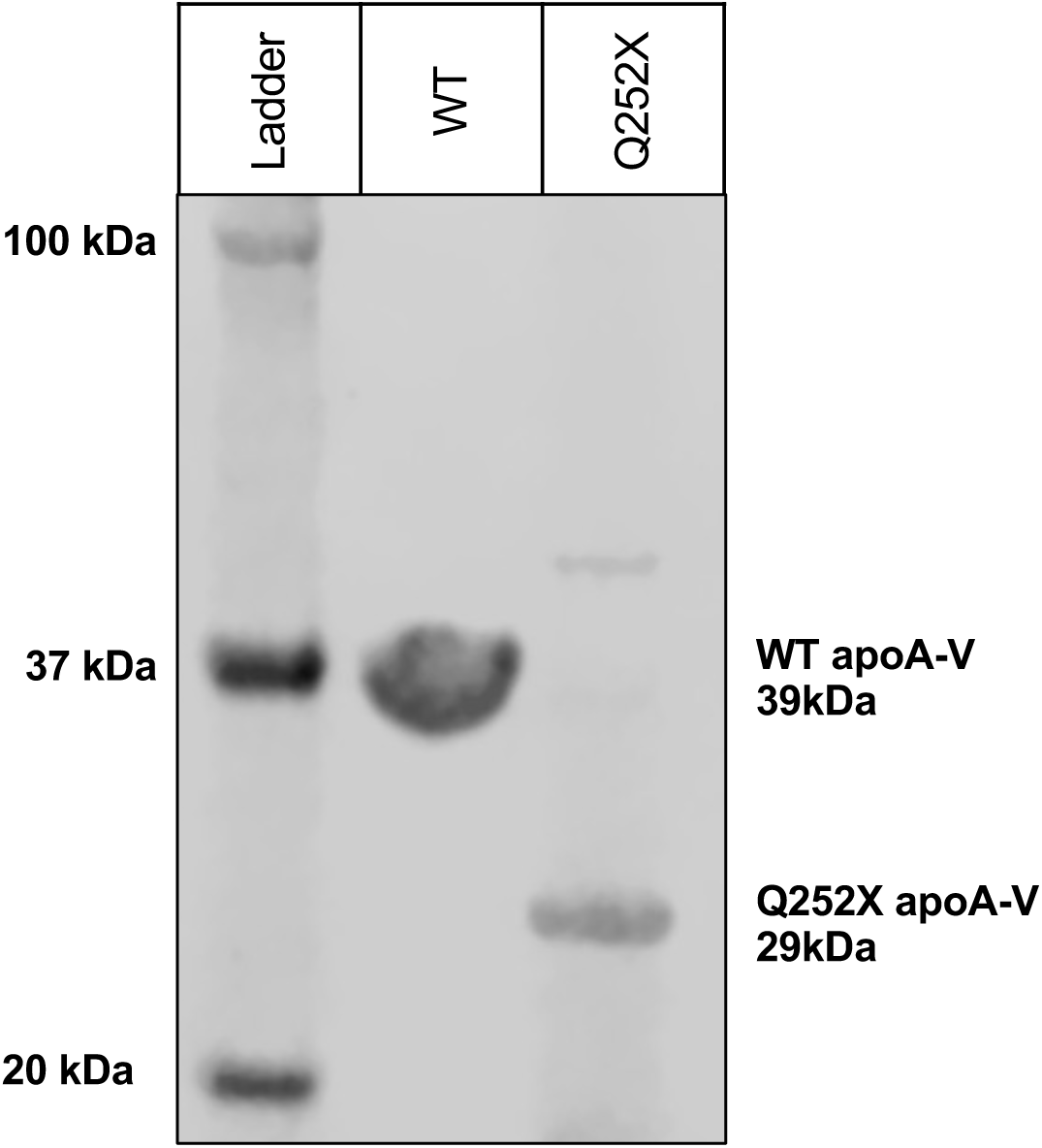
Plasma from AAV injected *apoa5* KO mice can be used to detect apoA-V Q252X. Recombinant WT and Q252X apoA-V protein was separated by SDS-PAGE. When diluted 1:100, plasma from *apoa5* KO mice injected with 3E12 vg Q252X h*APOA5* AAV can be used as primary antibody to detect WT apoA-V (39kDa) and apoA-V Q252X (predicted molecular weight 29 kDa).

**Figure S9.**
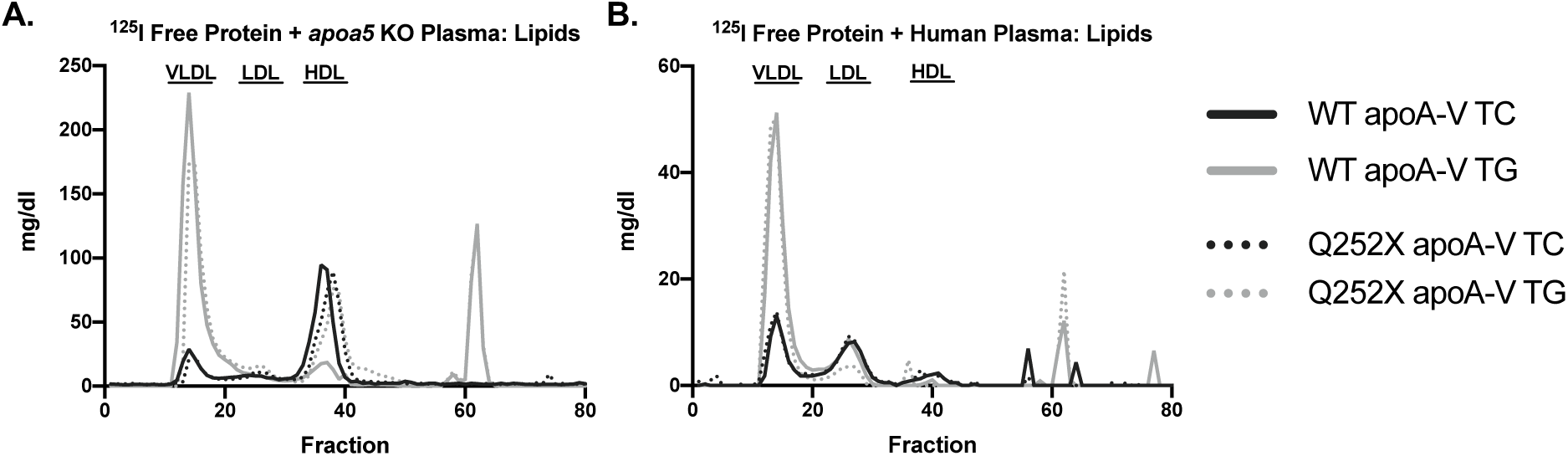
FPLC lipid profiles for assignment of lipoprotein elution ranges. ^125^I radiolabeled WT or Q252X apoA-V recombinant protein was incubated with plasma then separated by FPLC (Figure 5A**, 5B**). TG and cholesterol content in 100μl of each fraction for **A.** *apoa5* KO and **B.** human plasma. Points indicate single value. apoA-V, apolipoprotein A5; KO, knockout; TC, total cholesterol; TG, triglyceride.

**Figure S10.**
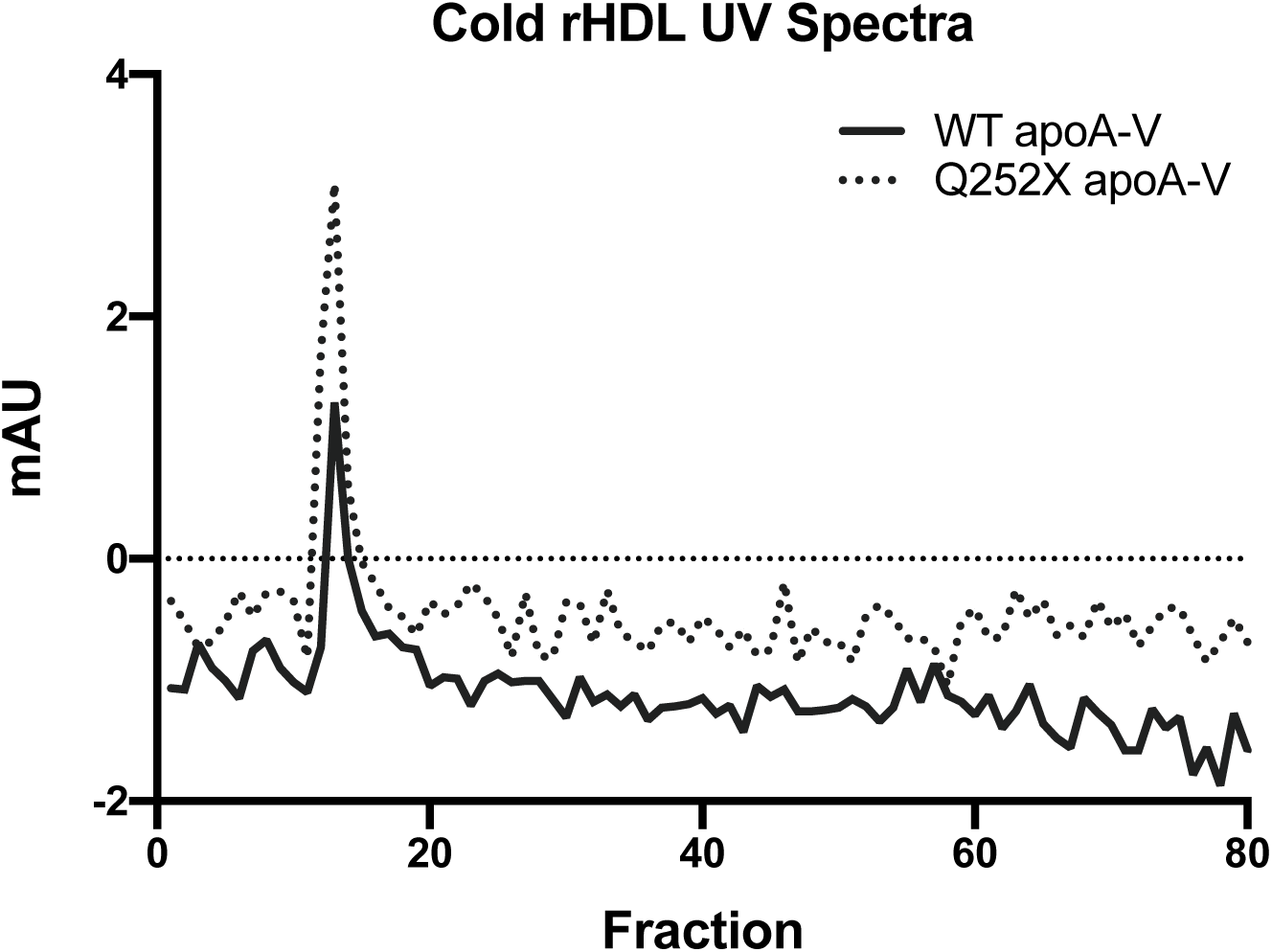
UV spectra of WT and Q252X apoA-V rHDL particles. Formulations were fractioned by FPLC and protein content in each fraction monitored by measuring the UV absorbance at wavelength 280nm. Points indicate single value. apoA-V, apolipoprotein A5; KO, knockout; UV, ultraviolet.

**Figure S11.**
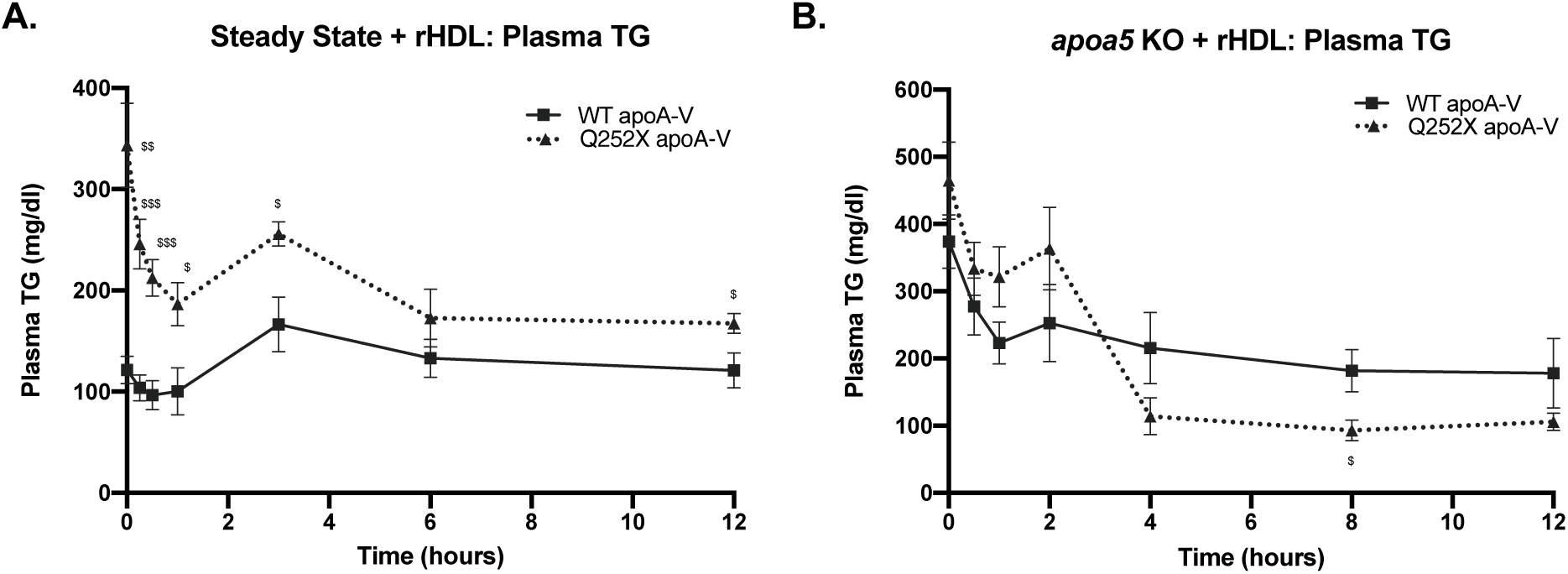
Plasma TG of *apoa5* KO mice administered apoA-V rHDL. **A.** *apoa5* KO mice expressing WT or Q252X apoA-V were administered 5μg WT or Q252X apoA-V synthetic HDL (rHDL) (N=5-6). **B.** *apoa5* KO mice were administered 20μg WT or Q252X apoA-V rHDL (N=7). apoA-V, apolipoprotein A5; KO, knockout; rHDL, synthetic HDL with recombinant apoA-V protein; TG, triglyceride.

## Notes

### Competing Interest Statement

Dr Rader is a consultant for Alnylam, Novartis, Pfizer, and Verve and has a grant from CSL

### Author Declarations

The University of Pennsylvania IRB

